# Interaction of surface type, temperature, and week of season on diagnosed concussion risk American football: Bayesian analysis of 8 Seasons of National Football League Data

**DOI:** 10.1101/2022.01.29.22270096

**Authors:** James M Smoliga, Sameer K. Deshpande, Zachary O. Binney

## Abstract

**Background:** Artificial turf fields and environmental conditions may influence sports concussion risk, but existing research is limited by uncontrolled confounding factors, limited sample size, and the assumption that risk factors are independent of one another. The purpose of this study was to examine how playing surface, time of season, and game temperature relate to diagnosed concussion risk in the National Football League (NFL).

**Methods:** This retrospective cohort study examined data from the 2012-2019 NFL regular season. Bayesian negative binomial regression models were fit to relate how playing surface, game temperature, and week of the season independently related to diagnosed concussion risk and any interactions among these factors.

**Results:** 1096 diagnosed concussions were identified in 1830 games. There was a >99% probability that concussion risk was reduced on grass surface (median Incidence rate ratio (IRR) = 0.78 [95% credible interval: 0.68, 0.89], >99% probability that concussion risk was lower at higher temperatures (IRR=0.85 [0.76,0.95] for each 7.9°C), and >91% probability that concussion risk increased with each week of the season (IRR=1.02 [1.00,1.04]). There was an >84% probability for a surface × temperature interaction (IRR=1.01 [0.96, 1.28]) and >75% probability for a surface × week interaction (IRR=1.02 [0.99, 1.05]).

**Conclusions:** Diagnosed concussion risk is increased on artificial turf compared to natural grass, and this is exacerbated in cold weather and, independently, later in the season. The complex interplay between these factors necessitates accounting for multiple factors and their interactions when investigating sports injury risk factors and devising mitigation methods.

## Introduction

Concerns regarding neurodegenerative diseases have prompted sports organizations to consider implementing policies and interventions aimed at reducing sport-related concussions.^1^ Identifying modifiable risk factors for concussions and repetitive head impacts in American football is particularly important,^2^ due to the potential risk of chronic traumatic encephalopathy.^3,4^

There has been considerable interest in examining the role of playing surfaces on brain injuries, and artificial turf fields are commonly marketed with claims of reducing the risk of concussions.^5-8^ Conversely, the National Football League’s (NFL) Players Association (NFLPA) claims that artificial turf is detrimental to player safety and long-term health,^9^ but there is currently limited evidence to support this claim specific to brain injuries. An analysis of the 2012-2013 seasons of National Football League (NFL) data found no association between playing surface and concussion risk.^2^ A 2019 systematic review and meta-analysis concluded that concussion risk was slightly decreased when contact sports (soccer, American football, and rugby union) were played on an artificial surface compared to natural grass.^10^ The two included studies for American football found no differences in concussion risk between one specific brand of artificial turf product and natural grass over three years of college football,^11^ and a greater risk of concussion on natural grass over five seasons of high school football.^12^

There are also within-season and temperature trends in injury rates in football and similar sports. Examination of four years of NFL data revealed a significant increase in concussion risk during the second half of the season, compared to the first half,^13^ and for games played in colder weather.^14^ Due to the timing of the season, which begins in late summer and ends in early January, and the primarily outdoor nature of the sport, these two factors are substantially correlated and should be considered together using methods to disentangle their associations. Surface may also interact with these variables. While biological factors, such as a reduced concussion threshold (i.e., the magnitude of injurious stimulus needed to produce concussion symptoms, which may be dependent on dynamic neuroanatomic and physiological factors and thus vary between and within athletes during a season), ^15-18^ could explain within-season changes in injury risk, it is also possible that within-season changes in the material properties of the playing surface (e.g., friction and shock absorption) could influence concussion risk.^19^ These may be due to wear/damage to the field^20-22^ or temperature-dependent effects,^23,24^ both of which may differ between playing surfaces. It is also possible that these apparent risk factors may not be purely physical/biological, but may also reflect bias in suspecting, reporting, or diagnosing concussions.

Playing surface, time of season, and game temperature all potentially influence concussion risk in football, but these factors are inter-related and existing analyses do not disentangle their potential confounding effects upon one another. Additionally, many analyses of these factors have divided temperature and week of season into categories,^2,13,14^ and it is possible that arbitrary categorization could lead to spurious associations for concussion risk.^25,26^ Thus, the objective of this study was to holistically examine how playing surface, game temperature, and week of season influenced diagnosed concussion risk in NFL games.

## Methods

### Study Design and Data Collection

We performed a retrospective cohort analysis of eight NFL seasons (2012-13 through 2019-20) of diagnosed concussion data from the PBS Frontline Concussion Watch^27^ and Football Outsiders^28^ injury databases. The Football Outsiders database includes data from all weekly NFL injury reports during this time period, including concussion and non-concussion injuries from weeks 1-16 of the regular season. To strengthen completeness, this concussion dataset was combined with data from the Frontline database, which provided an independently collected list of concussions incurred by an NFL player from 2012-2015. Previously identified erroneous data points within the Frontline data set^25^ were excluded. Injury report data from Week 17 are not available for teams that did not qualify for the playoffs, and therefore only the first 16 weeks of data (15 games per team) were analyzed, as is consistent with previous studies.^25^ We assumed that all concussions occurred in a game setting, unless 1) previous data from the Frontline database^25,27^ specified it occurred during a practice (in which case, it was excluded from analysis) or 2) if a concussion was listed for a player in the absence of him taking any snaps in games that week. Multiple analyses suggest that approximately 95% of diagnosed concussions in the NFL occur during games.^25,29^ However, this differs from youth, high school and college football, where concussions are more likely during practices,^30,31^ which is most likely due to the less frequent and more highly regulated nature of contact practices in the NFL during our study’s timeframe. We also assume that all of these concussions were diagnosed by appropriately trained medical personnel, and therefore consider all of the injuries in this analyses to be “diagnosed concussions” (heretofore, referred to as / used interchangeably with “concussions” unless otherwise specified).

#### Stadium and Playing Surface

Playing surface was categorized into natural grass, artificial turf, and hybrid (a surface which includes an artificial turf foundation, combined with natural grass, potential resulting in unique profile of material properties). Preliminary analysis revealed hybrid surfaces may be associated with a distinct difference in concussion risk compared to natural grass and artificial turf (SDC3: eTable 1, eFigure 1), but represented <5% of total games (Table 1). Thus, games played on hybrid surfaces (n=82) were excluded from analysis.

**TABLE 1.** Summary of NFL games included in the primary model. Note, games which were played in stadia with a retractable roof closed, but doors and windows open (n=8) are excluded from the table.

|  | **Indoor Games**<br>**(Concussions)** | **Outdoor Games**<br>**(Concussions)** | **Combined Games**<br>**(Concussions)** |
| --- | --- | --- | --- |
| **Artificial Turf** | 359 (218) | 441 (334) | 800 (552) |
| **Natural Grass** | 63 (25) | 967 (519) | 1030 (544) |
| **Hybrid Surface** | 0 (N/A) | 82 (52) | 82 (52) |
| **Combined**<br>**(Including Hybrid)** | 422 (243) | 1490 (905) | 1912 (1148) |
| **Combined**<br>**(Excluding Hybrid)** | 422 (243) | 1408 (853) | 1830 (1096) |

**Figure 1.**
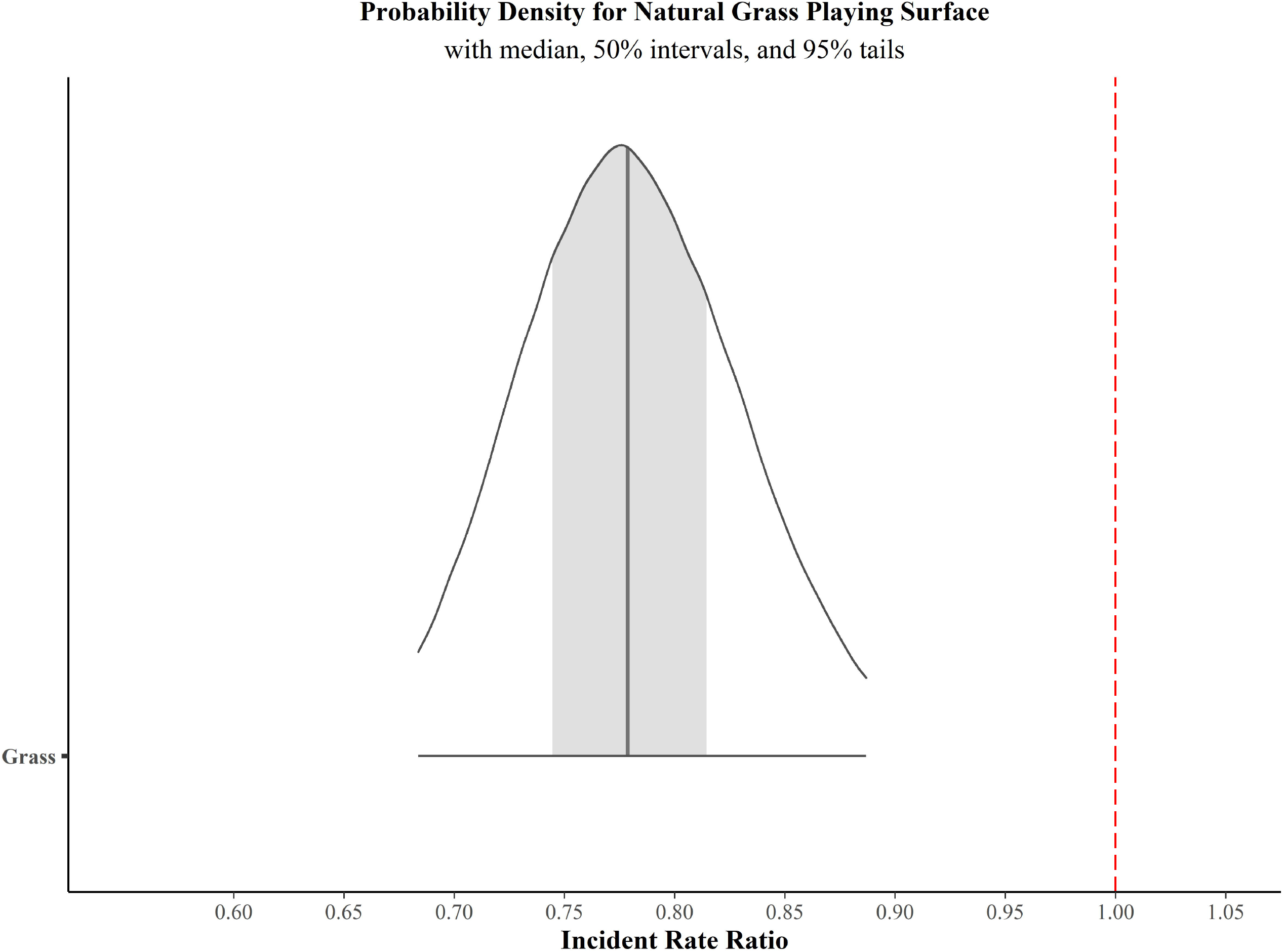
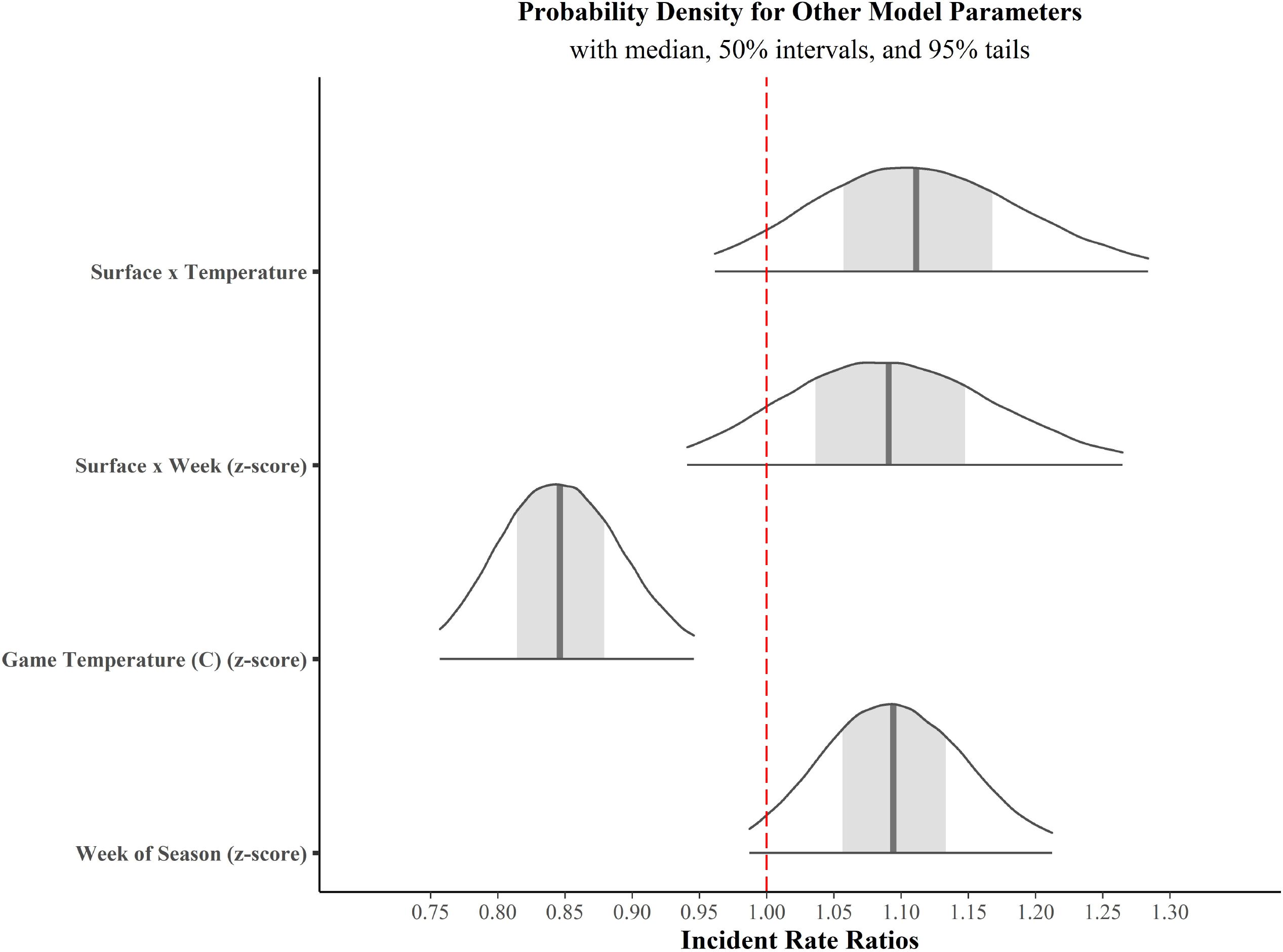
Probability distributions for all parameters primary model. All coefficients were exponentiated to compute Incidence Rate Ratio. For all figures, the vertical bar within each curve represents the median value for prior distribution, gray shading represents the 89% credible interval and the tails represent the 97% credible interval for each parameter coefficient. The intercept represents the concussion rate (number of concussions per game). The posterior distribution for the IRR of natural grass surface is below 1.0 (dotted red line, for frame of reference), representing a decreased risk for concussion. **Figure 1A. Probability distribution for natural grass playing surface**. The overall probability that concussion risk was reduced on natural grass compared to artificial turf was >98%, with a median incident rate ratio of 0.53 (∼47% reduced risk). However, positive values for two-way interactions (Figure 1C) suggest that the protective effect of natural grass may diminished with greater temperatures and later in the season. **Figure 1B. Probability distribution for game temperature, week of season, and the two-way interactions included in the model**. There was a high probability (>99%) that concussion risk was reduced at greater temperatures, and increased later in the season (>91%). Much of the density of the posterior distributions for week and the two-way interactions were >1.0, suggesting that these values may influence concussion risk, however cautious interpretation of these parameters may be warranted as their probabilities are a bit lower (Grass × Week: >75%; Grass × Temperature: >84%). If the two-way interactions are interpreted as “non-significant” and removed from the model, all main effect parameters increase to >99% probability of association with concussion risk (SDC3: eTable 3).

Aberrations from a team’s normal home stadium were accounted for in the dataset, including games played in international locations and games which were moved to a different stadium due to extenuating circumstances. Likewise, within-season and between-season changes in playing surface type within a stadium were accounted for (e.g., the Houston Texans played one game on natural grass before switching to artificial turf for the remainder of the 2015 season).

#### Game Temperature

The Pro-Football-Reference database^32^ provides official weather details for most games (n=1557 games). For outdoor games where any of this data was missing, temperature was retrieved from official NFL game reports and annual team media guides (n=247). If temperature was not provided in these sources (n=108 games), data were retrieved from WeatherUnderground^33^ from the zip code of the stadium at the nearest time point prior to kickoff (generally within one hour). The official NFL summary occasionally provided the temperature for indoor games (generally 20-22°C), and when this was not provided, it was assumed to be 20°C.

To determine game temperature in stadia with a retractable roof, the respective team’s media guide was examined to determine if the game was played under indoor conditions (i.e., roof closed, doors/windows closed) or outdoor conditions (i.e., roof open). If this information was not available in the media guide, Wikipedia game summaries, the official NFL summary, news records, and game video were used to determine whether the roof was open or closed. Games in which the doors/windows were open, but the roof was closed (n=8) were excluded from analyses, since game temperature was not certain.

## Data Analysis

### Primary Model

Data analysis was performed in R Studio v1.4.1106.^34^ The data set and source code are available as Supplemental Digital Content (SDC) 1 and 2, respectively. Preliminary analysis of the complete dataset revealed the number of concussions within a game is well-represented by a negative binomial distribution (eFigure 2). Thus, negative binomial regression models were developed to determine the relationship between risk factors and the number of concussions within a game. The number of concussions in a game served as the dependent variable. Week of season (henceforth referred to as “week”), playing surface (artificial turf vs. natural grass), and game temperature served as independent predictor variables.

**Figure 2.**
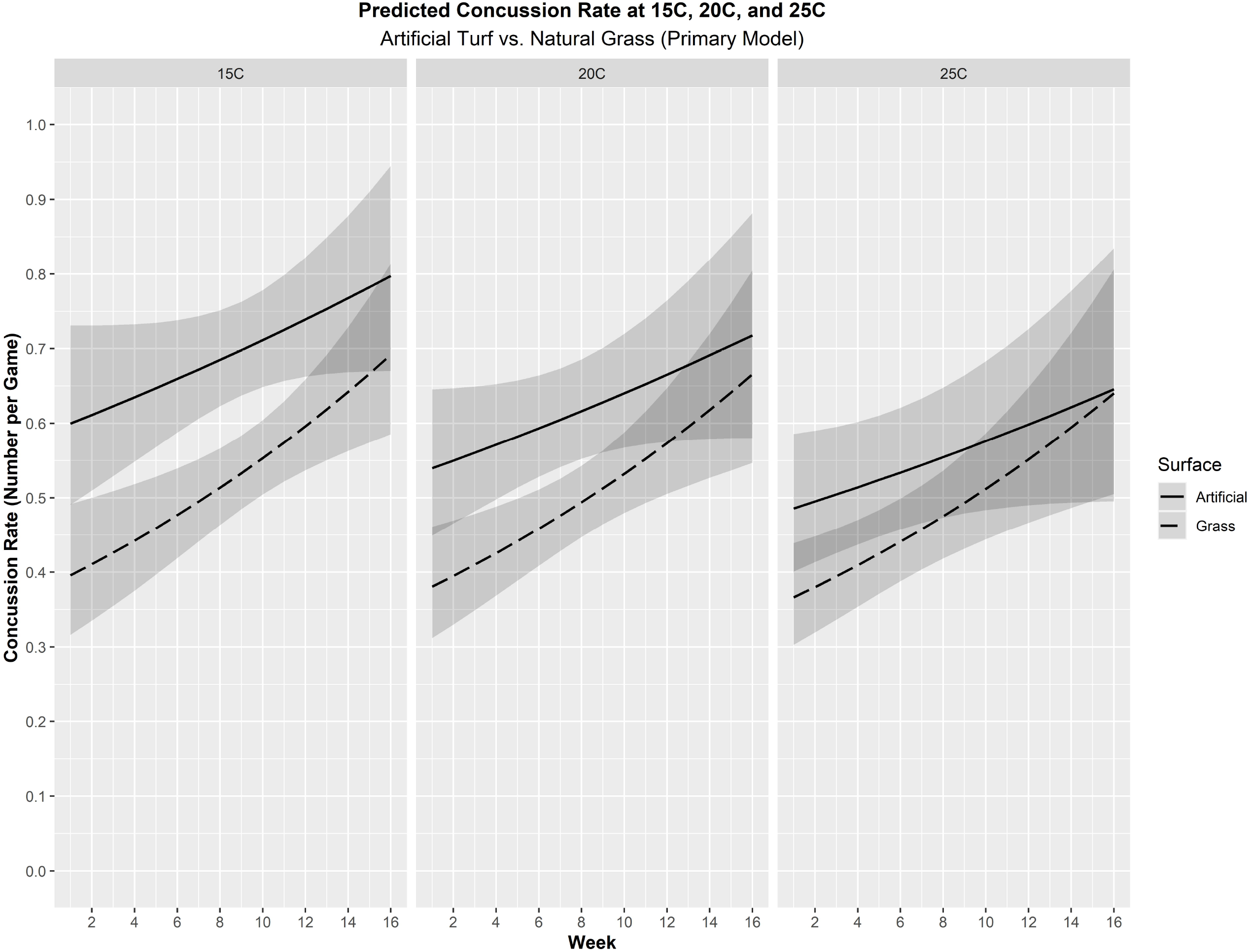
Marginal effects of concussion rate by week of season, at a fixed temperature of 20°C. The black spline (solid = artificial turf, dashed = grass) represents predicted concussion rate and grey shading represents the 95% credible intervals. These represent reasonable probable temperatures for outdoor games in temperate climates across the entire football season, and 20°C represents the approximate temperature for indoor games (see SDC3: eFigure 3A-C). At any given week of the season, the risk of concussion on artificial turf is magnified during colder temperatures.

As noted previously, surface wear occurs over the course of a season and this may differ between surface types. The material properties of playing surface also seem to be temperature dependent, and this may also vary by surface type. Thus, the two-way interactions of surface × week and surface × temperature were included in the model. We did not have any *a priori* justification to include the two-way interaction of week × temperature or the three-way interaction between all independent predictors in the model. However, we did fit these models in a supplementary analysis (SDC3: eTable 2A—B) and found similar results.

The model intercept represents the log of the average of number of concussions in a game played on artificial turf in the middle of the season (between weeks 8 and 9) and at an average temperature (17.1 degrees Celsius). Previous research suggests that over a 12 year span, the average number of concussions in an NFL was about 0.40.^35^ We therefore specified the prior distribution on the model intercept to be normal with mean -0.92 (*ln* 0.4 = -0.92) and with standard deviation 0.3. This choice represents a strong prior belief that in average conditions, there is often 0 or 1 concussions per game.

Previous research using Poisson regression models did not find any relationship between time of season (four categories of four weeks) or playing surface (natural grass versus artificial turf) across two NFL seasons.^2^ Thus, the priors for the model parameters corresponding to “week” and “playing surface” were set to 0 with a normal distribution with standard deviation 2.5. For simplicity, we specified identical normal priors for the remaining model parameters. Although these choices are somewhat informative, prior predictive checks^36^ revealed the priors on the parameters induced a fairly non-informative prior on the average number of concussions per game. To wit, the induced prior on average number of concussions per game placed 50% probability on the range [0.64, 4.39], 80% probability on the range [0.36, 20.23], 90% probability on the range [0.27, 69.14], and 95% probability on the range [0.21, 260.90]. Importantly, the induced prior does not concentrate on extremely small values near zero (which would suggest that there are essentially no concussions) or on implausibly large values (which would suggest that there are at least tens or hundreds of concussions per game). Instead, the induced prior covers a broad range, indicating that the prior regularization on the model parameters is not so strong as to bias our model parameters towards one set of conclusions.

We use the posterior mean as the point estimate of each model parameter and report the 95% central credible interval as a measure of uncertainty about these parameters. Since the posterior distribution is analytically intractable, we rely on Markov Chain Monte Carlo (MCMC) simulation to approximate these quantities. Our simulation was performed using the **rstanarm** package. We specifically ran 8 chains for 15,000 iterations each and discarded the first 5,000 iterations as warmup. and credible intervals for all model parameters.

We exponentiated each sample of the model parameters to compute the incidence rate ratio (IRR) of all of the predictive variables in our model. We performed several checks on the reasonableness of our fitted model (see SDC3).^36,37^

The median value and 95% central credible intervals were computed for the IRR of each parameter.^38^ We additionally computed the posterior probability that the IRR exceeded 1.0 (which would suggest a harmful effect), or was below 1.0 (which would suggest a beneficial effect).

### Sensitivity Analyses

To ensure the correlation between week and temperature did not cause collinearity problems for our primary model, we used separate models with novel control methods for each variable. We then compared the coefficients from these models to those of the primary model.

To determine if week of season was truly an independent predictor of concussion risk, we ran a model on only indoor games to control for temperature without including it in the model. This allowed for us to control for temperature (i.e., it would be similar for all games), without including it as a model parameter, and determine if week was still an independent predictor of concussion. We ran a similar model using both indoor games and outdoor games played within a narrow temperature range (17.8 to 25.6°C).

To determine if temperature was an independent predictor, we ran a model on only games during weeks 13-16 without a term for week in the model. We would not expect a major change in concussion risk from week alone during this short 4-week span, but games during this time have a wide range of temperatures. Thus, this late season model would allow for us to determine if temperature was still a predictor of concussion within this short time span.

## Results

### Games Included

There were 1920 games played during the first 16 weeks of the 2012-2019 seasons (Table 1). Of these, a total of 1830 games met the inclusion criteria described in the methods and were included in the primary model. Median temperature was similar between surfaces (∼20°C for both), but temperature distribution varied by playing surface and week of season. Games played on natural grass (n=1030) followed a left-skewed distribution, whereas games played on artificial turf (n=800) had a spike at 20°C (n=359 indoor games at 20°C, with n=441 outdoor games with median 13.9°C) (eFigure 3A-B). Median temperature decreased over the course of the season (eFigure 3C).

### Model Results

#### Model diagnostics and fit

Our diagnostics did not reveal problems with convergence or mixing or the simulated Markov Chains: all of the R-hat statistics were close to 1.0 (consistent with convergence to the stationary distribution) and visual inspection of the traceplots did not reveal issues with mixing. Additional checks revealed that there was no single game exerting undue influence on our posterior distribution. Detailed discussion of our model diagnostics are available in the SDC3.

#### Model parameters

A summary of model parameters are presented in Table 2 and probability density plots for the posterior distribution with IRR credible intervals for each parameter are provided in Figure 1. Grass playing surface (>99% probability), early season games (>91% probability), and higher temperature (>99% probability) games are all independently associated with a decreased risk of diagnosed concussion.

**Table 2.**
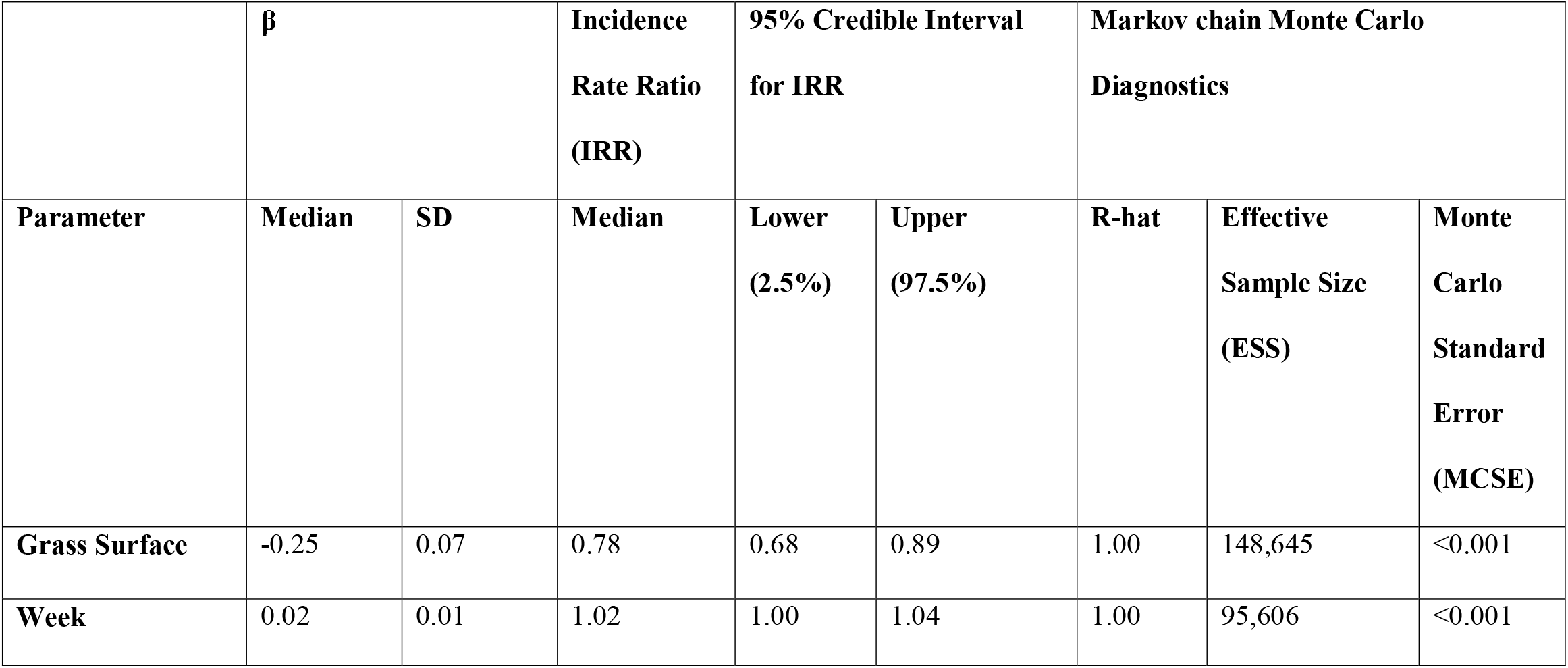
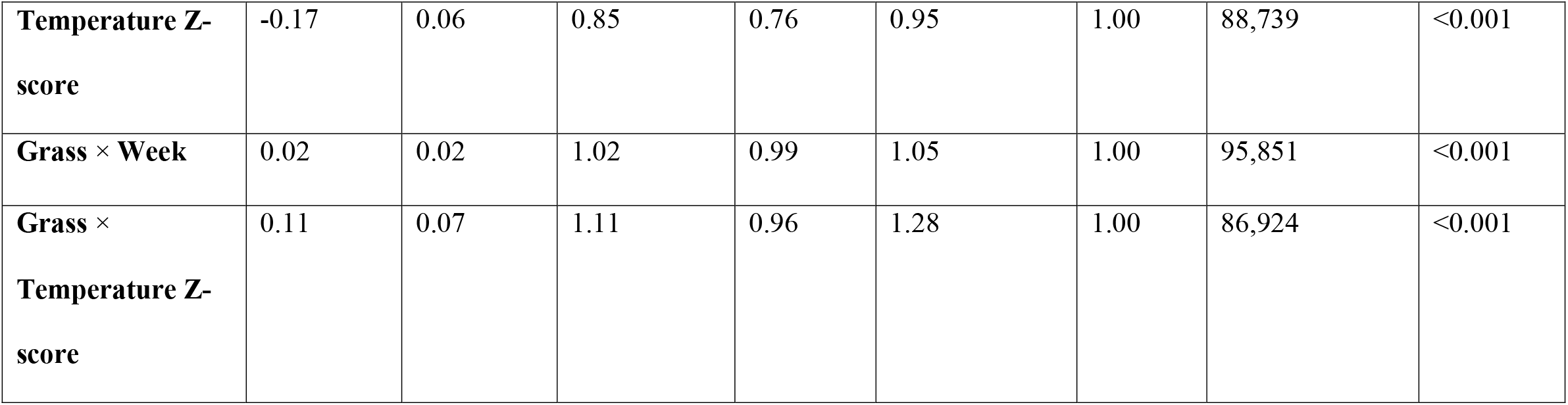
Summary of parameters from the primary negative binomial regression model, including relevant two-way interaction terms (n=1830 games). Grass surface was associated with a reduced risk of concussion compared to artificial surface (>99% probability). Concussion risk was also reduced with greater temperatures (>99% probability) and increased with each week of the season (>91% probability). There is reasonable probability (>75%) that each of the two-way interaction terms affect concussion risk. If the two-way interaction terms are interpreted as “no effect” and removed from the model, all three main effects have a >99% probability of influencing concussion risk (SDC3: eTable 3, eFigures 6A-B and 7A-B). (Note, the lower bound credible interval for Week is reported as 1.00, but is 0.997, so the 95% credible interval does cross 1.0.)

| | $\beta$ | | Incidence<br>Rate Ratio<br>(IRR) | 95% Credible Interval<br>for IRR | | Markov chain Monte Carlo<br>Diagnostics | | |
| --- | --- | --- | --- | --- | --- | --- | --- | --- |
| Parameter | Median | SD | Median | Lower<br>(2.5%) | Upper<br>(97.5%) | R-hat | Effective<br>Sample Size<br>(ESS) | Monte<br>Carlo<br>Standard<br>Error<br>(MCSE) |
| Grass Surface | -0.25 | 0.07 | 0.78 | 0.68 | 0.89 | 1.00 | 148,645 | <0.001 |
| Week | 0.02 | 0.01 | 1.02 | 1.00 | 1.04 | 1.00 | 95,606 | <0.001 |
| **Temperature Z-score** | -0.17 | 0.06 | 0.85 | 0.76 | 0.95 | 1.00 | 88,739 | <0.001 |
| **Grass × Week** | 0.02 | 0.02 | 1.02 | 0.99 | 1.05 | 1.00 | 95,851 | <0.001 |
| **Grass ×**<br>**Temperature Z-score** | 0.11 | 0.07 | 1.11 | 0.96 | 1.28 | 1.00 | 86,924 | <0.001 |

The point estimate for grass surface IRR was 0.78, meaning that for mid-season games (weeks 8 and 9) played at the average temperature for the dataset (∼17°C), the risk of concussion was ∼22% lower on grass than on artificial turf. The 95% credible intervals (0.68, 0.89) indicated the true effect is likely is between a 11-32% reduction on grass compared to artificial turf.

There was >75% probability that a two-way interactions of surface × week influenced concussion risk (Figure 1B), with an estimated one-week IRR of 1.02 (1.00, 1.04) on artificial turf and 1.04 (1.01, 1.06) on grass. In other words, for each one-week change, there was a ∼2% increased risk of concussions on artificial turf and ∼4% increased risk on grass, when temperature was held constant at the average for the dataset. There was >84% probability that the two-way interaction of surface × temperature influenced risk. The point estimates for temperature IRR were 0.84 (0.76, 0.95) on artificial turf and 0.94 (0.86, 1.03) on grass. This translates to a ∼16% reduced/increased risk of concussion on artificial turf and a ∼6% reduced/increased risk of concussion on natural grass, for each one standard deviation (corresponding to 7.9°C) increase/decrease in temperature during the middle of the season.

The results indicate the protective association of grass surface compared to artificial turf was somewhat attenuated in warmer conditions and late-season games (Figures 2 and 3). While the risk of concussion increases over the course of the season regardless of surface, this weekly increase in risk seems to be greater on natural grass than it does on artificial turf when temperature is held constant (Figure 2). The risk of concussion seems to increase substantially on artificial turf in colder temperatures, whereas cold temperatures do not seem to exacerbate the risk of concussion on natural grass as much holding week constant (Figure 3).

**Figure 3.**
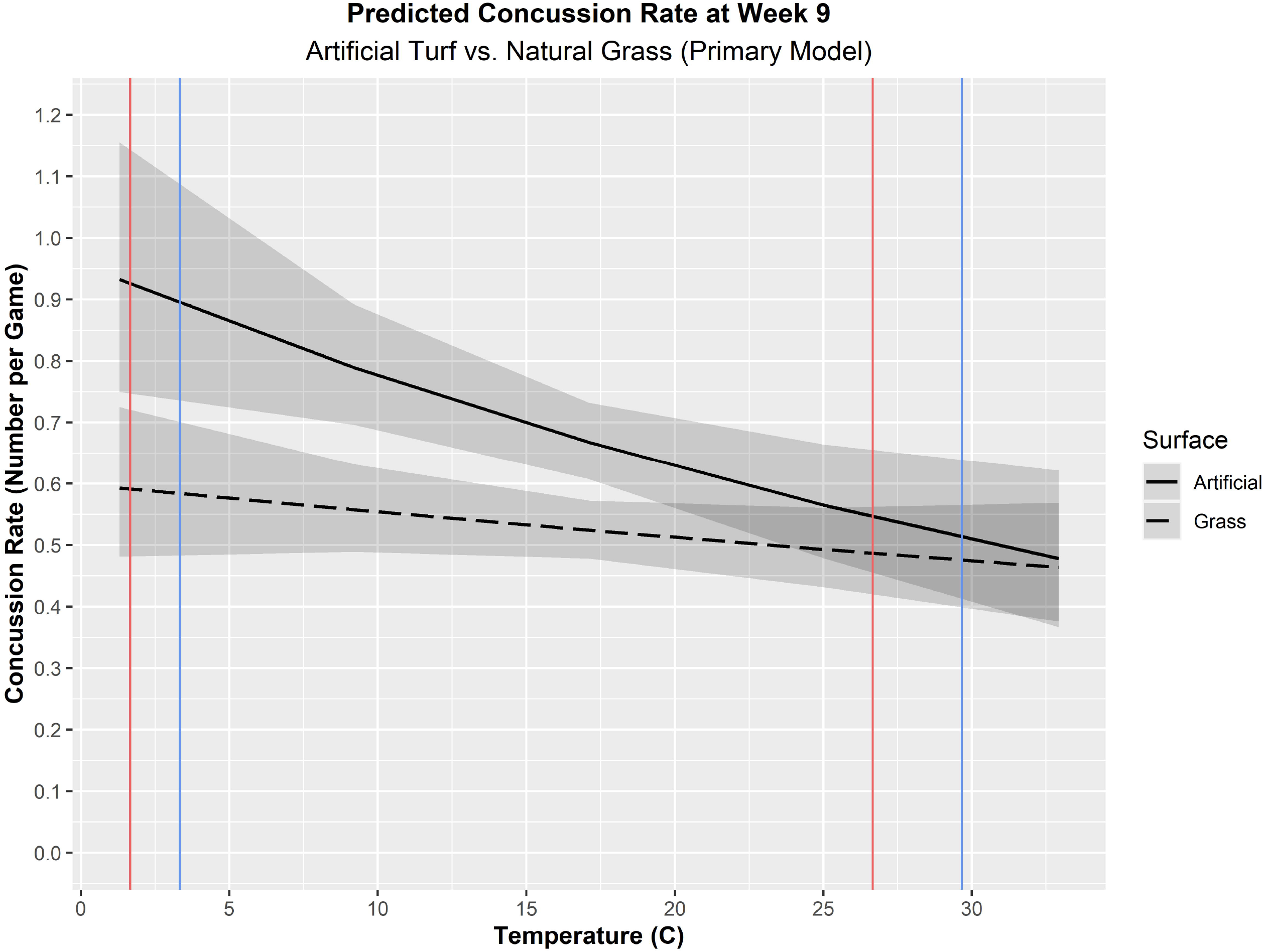
Marginal effects of concussion rate by temperature, fixed to Week 9 of the NFL season. The black spline (solid = artificial turf, dashed = grass) represents predicted concussion rate and grey shading represents the 95% credible intervals. Week 9 is utilized for visualization because it represents a time of the season with a wide range of game temperatures. The red and blue vertical lines represent the temperature extremes observed on at Week 9 within our dataset for artificial turf (n=44 games, mean temperature = 13.4°C) and natural grass (n=57 games, mean temperature = 17.4°C), respectively. For games as cold as 1.5°C, our model is quite certain that there will be more concussions on artificial turf than grass. At the other end of the temperature spectrum, however, our model is much more uncertain, as indicated by the substantial overlap in the credible intervals at temperatures above 20°C. This overlap is to be expected due to low sample size at higher temperatures at Week 9, typically falls in the early part of November. For Week 9 games, the average temperature in outdoor stadiums (n=88 games, artificial turf and natural grass combined) is 15.0°C (standard deviation = 6.8°C).

Though the two-way interactions were likely associated with concussion risk (>75% probability), we also ran the models without these interactions to see if the results for the main effects remained stable. When this main effects model is used, all three independent predictors have a >99% probability (SDC3: eTable 3A, eFigures 6A-C) and produced nearly identical point estimates to the primary model (SDC3: eTable 3B).

#### Diagnosed concussion rates per game

For an average-temperature (∼17°C), mid-season game (Week 8-9), the number of concussion predicted per game was 0.51 on natural grass and 0.66 on artificial turf. Early season (Week 1) warm weather (25°C) games have a concussion rate of 0.37 per game on natural grass and 0.48 per game on artificial turf. Late season (Week 16) cold weather (5°C) games have concussion rates of 0.72 and 0.96 per game, on the respective surfaces.

#### Sensitivity Analyses

Models investigating only indoor games or indoor and outdoor games in a moderate temperature range found similar IRRs for week and its interaction with surface as the main model (Table 2 and SDC3: eTable 4B). A model investigating only games in weeks 13-16 found similar IRRs for temperature and its interaction with surface as the main model (Table 2 and SDC3: eTable 5B). This confirmed both as independent predictors.

## Discussion

To our knowledge, this is the first study to comprehensively examine how the interaction of surface, temperature, and time in season factors interact to influence concussion risk and the first to use a Bayesian approach to evaluating concussion risk. We found that the risk of diagnosed concussion is reduced on natural grass playing surfaces, increased later in the season, and increased in colder temperatures. While all of these factors independently modify diagnosed concussion risk, week and temperature both interact with the effect of playing surface. We posit that these factors modify the effect of playing surface (due to changes in surface material properties, described in *Explanatory Mechanisms* section), but it is also possible that playing surface modifies the effects of week and temperature. These findings provide compelling evidence that artificial turf is associated with greater concussion risk in the NFL, and support the NFL Player’s Association’s advocacy for transitioning to natural grass fields to improve player safety.^9^ Additionally, these results suggest that extending the NFL season will likely increase concussion risk.

The large sample size combined with the Bayesian negative binomial regression model provide us with a high-level of confidence in our results, which conflict with some existing literature with smaller sample sizes.^2^ In an industry-funded study, Meyers found a slightly greater risk of concussion on natural grass compared to one specific manufacturer’s (FieldTurf) artificial turf over three seasons of college football.^11^ We ran a separate analysis comparing natural grass surfaces to FieldTurf surfaces, and found natural grass was still associated with lower concussion risk (SDC3: eTables 6A-B, Figures 8A-B). Discrepancies may be attributable to greater sample size in our study, differences between college versus professional football, or differences in time period studied. The evidence that week and temperature interact with surface effects may also explain conflicting findings within the literature regarding the role of playing surface in athletic injuries.

### Explanatory Mechanisms

The most likely explanation for lower concussion risk on natural grass compared to artificial turf is rooted in the biomechanics of helmet-to-ground impacts. Video review of injuries across five NFL seasons (1996-2001), suggests helmet-to-ground impacts account for 16%^39^ to 22%^40^ of concussions. A more recent study across two seasons (2015-2016) found 18% of concussion with a clear source of impact were due to this mechanism.^41^ The biomechanics of helmet-to-ground impacts are substantially different than player-to-player impacts, and football helmets are not tested (or designed) for these types of impacts.^42-44^ These impacts are potentially more severe, due to unusually high angular velocities and accelerations, and the potential for rebounds (both of the head within the helmet, and with the helmet and head combined).^43^

Grass tears in relation to translational force, whereas artificial turf does not – which allows the latter to have greater force transmission between the player and the surface.^45,46^ Increased friction between the helmet and surface could lead to greater torque placed about the head upon a helmet-ground interaction.^43^ Indeed, laboratory reconstruction of helmet-to-ground impacts has revealed the severity of impact is highly sensitive to the compliance and frictional properties of the surface.^42^ Reconstruction of 10 representative impacts causing concussion from NFL games demonstrated greater severity index and head injury criterion for all helmet-to-ground impacts on artificial turf (n=2) compared to helmet-to-helmet impacts (n=8).^47^ If our results are indeed driven by surface materials, this has implications for other athletes, since head-to-ground impacts account for 5-20% of concussions across various sports.^48-53^

Week of season and game temperature were independently found to influence concussion risk, which are also likely rooted in helmet impact biomechanics. Both surfaces degrade over the course of the season,^20,21^ which can influence friction and energy absorption.^22,54^ Likewise, surface temperature influences shock absorption properties of artificial turf,^23,24^ and soil hardness and grass quality of soccer fields (moisture dependent, with the greatest effect in winter months).^54^ Artificial surface temperature is also highly influenced by solar radiation,^55^ which varies with season. As demonstrated by the interaction effects (Figures 1C, 2A-B), there is a reasonably high probability that natural grass fields may also be prone to greater degradation within a season compared to artificial turf (e.g., soil compaction, significant decrease in grass density, etc.),^54,56^ and lose some of it apparent protective effect later in the season.

Various biological factors could independently account for increased concussion risk later in the season. It has been theorized that prior history of repeated sub-concussive head impacts could change one’s biological threshold for a concussion,^57^ and there is mixed evidence to support this.^15-18,58-61^ Additionally, cross-sectional data suggests that brain volume changes seasonally in the general population,^62^ and this could theoretically influence concussion risk over the course of a season, regardless of game conditions or cumulative impact exposure.

It is also possible that these risk factors also influence an athlete’s perception of concussion risk, concussion signs/symptoms, and/or voluntary reporting of concussion-related concerns. These factors may also influence medical personnel’s vigilance in suspecting a concussion and/or accurately diagnosing it. For instance, if players or medical staff believe that artificial turf increases concussion risk, it is possible that they are more likely to suspect/report/diagnose a concussion after a helmet-to-ground impact on artificial turf than a similar impact on natural grass. NFL players have expressed concerns about playing surface,^9^ and elite soccer players also perceive surface influences injury risk,^63^ and some even believe this is temperature-dependent.^64^ However, it is unknown how these perceptions influence self-recognition of injury symptoms and reporting to medical staff. It is also possible that athletes may be more or less likely to report symptoms at different points of the season and may also be influenced by high-stakes competitive outcomes.^65^ Indeed, NFL players diagnosed with a concussion are more likely to face salary reductions and be released from their team.^66^

### Limitations

Concussions are under-diagnosed across various sports and proficiency levels, so it is possible that some of these injuries were missed and our analysis cannot provide any insight on those. Players are tagged by medical staff to be evaluated for concussion based on high-risk situations and visible clinical signs (e.g., ataxia, loss of consciousness, slow to get up),^67^ but these are not as common as headache or dizziness (reported by ∼55% and ∼41% of documented concussions in NFL players, respectively).^35^ Analysis of the 2017 NFL season video footage revealed ∼26 of players diagnosed with a concussions did not exhibit visible signs, and thus required more thorough medical evaluation.^68^ This means that concussion recognition / diagnosis is highly (but not entirely) dependent on honest reporting from players. Survey research has revealed ∼23% of collegiate football did not intend to report concussion symptoms (with another 9.6% uncertain if they would),^69^ but concussion non-disclosure data from modern-day NFL players are not available. Thus, the data used in this study (and much of the sports-related concussion epidemiology literature) only represent documented/diagnosed concussions. Therefore, we are confident that our model identifies risk factors for diagnosed concussions, but it is unknown if it is representative of all concussions (i.e., documented and undocumented). This would likely be the case if these risk factors were purely physical/biological, but it is possible that concussion reporting / diagnosis is biased in either direction by some of these risk factors (as described in the previous section).

There is also considerable heterogeneity within the broad categories of artificial turf and natural grass,^46^ and we did not attempt to differentiate between different them (with the exception of Field Turf, SDC3: eTable 6A-B). In other sports, certain species/cultivars may be associated with differing risks of musculoskeletal injuries independent of ground hardness (attributable to differences in thatch),^70,71^ though not concussions.^71^ Surface moisture can also influence surface material properties.^72^ One could attempt to account for this by including weather in analysis, however, pre-existing precipitation influence the surface even if game-time weather as dry and weather can vary considerably within a game. We were also unable to quantify field “wear” given different environmental conditions (e.g., sunlight) and non-game activity (e.g.., concerts, setup and maintenance),^73^ and these may vary between stadia.

Given the increased risk of concussion later in the season and at colder temperatures, we suspect that concussion risk would be greatest during the playoff season. However, accurate injury reports are not available for teams which fail to advance to the next round of playoffs (or after the championship game), which precludes the possibility of a valid analysis of concussion risk during this time.

## Conclusions

Eight years of data demonstrated that diagnosed concussion risk is substantially increased in NFL games played on artificial turf. While concussion risk also increases with cold weather regardless of playing surface, the risk is particularly amplified on artificial turf. There is also an increased risk of concussion later in the season, even when temperature is controlled for, but it remains uncertain if is this is due to biological factors, field degradation, or a combination. These data suggest that extending the NFL season may provide greater health risk to players, and that player concerns over the use of artificial turf are warranted. Further research is necessary to determine if these findings extend to non-professional levels of American football, and other team sports.

## Supporting information

Supplemental Methods and Results

## Data Availability

All data produced in the present study will be available upon acceptance into a peer-review journal.

