## Supplemental Methods and Results for "Interaction of surface type, temperature, and week of season on diagnosed concussion risk American football: Bayesian analysis of 8 Seasons of National Football League Data"

**Supplemental Digital Content**

**Table of Contents**

**eMethods**

**eResults**

**eTable 1.** Preliminary analysis utilizing a main effects negative binomial regression model, including games played on artificial turf, natural grass, and hybrid surfaces (n=1912 games).

**eTable 2**

**eTable 2A.** Three-way interaction negative binomial regression model (n=1830 games).

**eTable 2B.** Two-way interaction (all) negative binomial regression model (n=1830 games).

**eTable 2C.** Two-way interaction (all) negative binomial regression model (n=1830 games), with relaxed prior (center = 0, standard deviation = 0.25)

**eTable 2D.** Two-way interaction (all) negative binomial regression model (n=1830 games), with relaxed prior (center = 0, standard deviation = 1.0)

**eTable 3.**

**3A.** Main effects negative binomial regression model.

**3B.** Point estimates from selected game conditions using main effects model.

**eTable 4.** Negative binomial regression models using a subset of data at relatively fixed temperature.

**4A** Only indoor games on artificial turf (n=359).

**4B** All games (natural grass + artificial turf) played at a temperature 17.8 to 25.6^o^C (n=827) – interactions included

**4C** All games (natural grass + artificial turf) played at a temperature 17.8 to 25.6^o^C (n=827) – main effects-only.

**eTable 5.** Negative binomial regression models using a subset of late season games (weeks 13-16)

**5A.** Model with two-way interactions.

**5B.** Main effects-only model.

**eTable 6.** Summary of regression parameters when natural grass was compared to Field Turf brand artificial surface**.**

**6A.** Relevant two-way interaction terms included.

**6B.** Main effects-only model.

**eFigure 1.** Posterior distribution of surface parameter estimates in the preliminary main effects model, including games played on hybrid surfaces.

**eFigure 2.** Histogram of the number of concussions per game across 8 NFL seasons.

**eFigure 3.** Distribution of temperature by surface and by week.

**3A.** All games (indoor + outdoor) on natural grass.

**3B.** All games (indoor + outdoor) on artificial turf.

**3C.** Comparison of Temperature by Week of NFL Season

**eFigure 4.** Monte Carlo Markov Chain (MCMC) trace plots for each parameter in the model.

**eFigure 5.** Observed vs. model-predicted counts of concussion across NFL games.

**5A.** Natural grass and artificial turf combined.

**5B.** Natural grass and artificial turf separated.

**eFigure 6.** Marginal effects of concussion rate by week of season at fixed temperature of 15^o^C, 20^o^C, and 25^o^C, using main effects only model.

**eFigure 7.** Marginal effects of concussion rate by week of season at fixed week of the season (Week 9), using main effects only model.

**eFigure 8.** Marginal effects of concussion rate comparing natural grass to FieldTurf artificial surfaces using main effects-only model

**8A.** By week of season at fixed temperatures, using main effects only model

**8B.** Fixed to Week 9 of the NFL season

**eMethods**

**Relationship between week and game temperature**

To determine if week and temperature were correlated to each other, Spearman’s correlations were computed. This was performed for all games (indoor + outdoor), as well as outdoor-only games.

**Comparison of game temperature by surface**

To determine if temperature was different between natural grass and artificial turf, an analysis of variance (ANOVA) was performed, using playing surface as the independent predictor variable. This analysis was performed for all games (indoor + outdoor), as well as outdoor-only games.

**Convergence and model fitting diagnostics**

We computed the Rhat statistic and effective sample size for the simulated samples of each model parameter. Effective sample size was >10,000 for each model parameter, and R-hat was ~1.000 for each parameter. Post-warmup trace plots demonstrated convergence to a common distribution for all chains, with all chains achieving stationarity (eFigure 4).38

We additionally performed a leave-one-out cross-validation analysis to assess whether or not our model overfit the observed data. This analysis was performed using the LOO package, ^1^ which reports the Pareto k diagnostic that can assess severe model misspecification. Evaluation of the Pareto smoothed importance sampling revealed all k values < 0.5, suggesting there were no games points which had undue influence on the posterior distribution. A power density overlay plot revealed the model was an excellent fit (eFigure 5A-B).

Graphical posterior predictive checks were performed to determine if the model was a good fit. Observed versus model-predicted number of concussions from 1000 simulations using the posterior predictive distributions were plotted and visually assessed.

**Models controlling for game temperature**

The primary model suggested that Week and Game Temperature both independently contributed to concussion risk. However, Week and Temperature are correlated to one another for outdoor games (n=1408, Spearman’s ρ = -0.582, p<0.001), and also for the whole data set when indoor games are included (-0.486, p<0.001). Thus, models controlling for game temperature were developed to determine whether week was confounded by game temperature. This was achieved by analyzing two subset of data:

1) games played indoors on artificial turf (n=359), and

2) all indoor and outdoor games played at 17.8 to 25.6^o^C (64 to 78^o^F, representative of the extremes of the temperature range at which indoor games are played at, n=827). The interaction between Surface × Week was originally included in this model, but was later removed due to its weak effect.

**Models controlling for week**

Along with the above models controlling for temperature, we attempted to confirm that temperature influenced concussion risk, independent of week. To do this, we ran our model on a subset of data consisting only of Weeks 13-16. This range of weeks was chosen since there is considerably variation in temperature during this period of the NFL season. While week could still be associated with concussion risk, we expected that the effect would be trivial during this short, 4-week time span, and thus any difference in concussion risk would be more attributable to surface and temperature. Thus, we originally included the main effect of week, and the two-way interactions of Surface × Week and Surface × Temperature in the model, but later removed the interactions due to weak effects.

**Analysis of limited FieldTurf artificial surfaces**

Previous research reported that the FieldTurf brand of artificial surface was associated with a slightly, albeit non-significant, reduced concussion rate in American football compared to artificial grass over three seasons, so we performed a sub-analysis using only this type of surface.

**eResults**

**Relationship between week and game temperature**

Week and temperature are correlated to one another for outdoor games (n=1408, Spearman’s ρ = -0.582, p<0.001), and also when indoor games are included (-0.486, p<0.001).

**Comparison of game temperature by surface**

The distribution of game temperatures differed between games played on natural grass compared to those played on artificial turf (eFigure 3), in part due to the large number of indoor games played on the artificial turf. Game temperature was significantly (p<0.001) warmer on natural grass fields (63.8 [62.7, 64.7]) than artificial surfaces (55.8 [54.3, 57.2]). This holds true when indoor games are included (64.0 [63.1, 64.9] vs. 61.3 [60.3, 62.3).

**Primary model diagnostic checks**

Evaluation of the Bayesian posterior distribution diagnostic data and leave one out validation parameters revealed no abnormalities. Effective sample size was >10,000 for each model parameter, and R-hat was ~1.000 for each parameter. Post-warmup trace plots demonstrated convergence to a common distribution for all chains, with all chains achieving stationarity (eFigure 4).38 Evaluation of the Pareto smoothed importance sampling revealed all k values < 0.5, suggesting there were no games points which had undue influence on the posterior distribution. A power density overlay plot revealed the primary model was an excellent fit (eFigure 5A-B).

**Models controlling for game temperature**

Results from these models are presented in eTables 4A-C. Regardless of the model used, it was highly probable (90-99%) that week was associated with concussion risk at games played at a relatively fixed temperature (17.8 to 25.6^o^C). This suggests that week of season is an independent predictor of concussion, separate from that of game temperature.

**Models controlling for week**

Results from these models are presented in eTable 5A-B. There was a 92% probability that temperature was associated with concussion risk, and a 98% that surface was associated with concussion risk. In this model, with only 4 weeks of data, there was <50% probability that week was associated with concussion risk. This suggests that game temperature is an independent predictor of concussion, separate from that of week of the season.

**Analysis of limited FieldTurf artificial surfaces**

The results were generally consistent with our results for the full sample (i.e., all artificial surface manufacturers), such that natural grass surface was associated with a *reduced* risk of concussion compared to FieldTurf products. The two-way interaction model indicated that Surface × Week had a minimal effect on concussion risk (eTable 6A) and therefore this interaction term was removed from the model. The new model, with only one two-way interaction (Surface × Temperature) was a better fit (eTable 6B).

Regardless of the model used, marginal effects plots suggest that natural grass is associated with a reduced risk of concussion compared to FieldTurf, which is magnified at cold temperatures and later in the season. However, there is considerable overlap in the 95% credibility interval at warmer and colder temperatures, and this is likely attributable to smaller sample sizes compared to the primary analysis (which includes all manufacturers of artificial turf). Meyers had a sample size of 465 games (n=230 on FieldTurf and n=235 on natural grass). Meyers dichotomized temperature into “hot” (>70^o^F) and “cold” (≤69^o^F) and did find significantly more injuries on FieldTurf during “cold” weather, but the number of games at each temperature was not specified, and it was unclear if this held true specifically for concussions.

**eTable 1. Preliminary analysis utilizing a main effects negative binomial regression model, including games played on artificial turf, natural grass, and hybrid surfaces (n=1912 games).** Median values of Incidence Rate Ratio (IRR) <1.0 suggest that natural grass and hybrid surfaces were both associated with a lower risk of concussion compared to artificial turf. However, the posterior distributions differed substantially between surfaces, with substantially narrower credible intervals for natural grass (see eFigure 1). The probability that hybrid surface was associated with reduced concussion risk was 69%, whereas natural grass had a 99% probability of reduced concussion risk.

|  | **β** | | **Incidence Rate Ratio (IRR)** | **95% Credible Interval for IRR** | | **MCMC Diagnostics** | | |
| --- | --- | --- | --- | --- | --- | --- | --- | --- |
| **Parameter** | **Median** | **SD** | **Median** | **Lower (2.5%)** | **Upper (97.5%)** | **R-hat** | **ESS** | **MCSE** |
| **Surface** |  |  |  |  |  |  |  |  |
| ***Grass*** | -0.25 | 0.06 | 0.78 | 0.68 | 0.88 | 1.00 | 164,229 | <0.001 |
| ***Hybrid*** | -0.16 | 0.16 | 0.85 | 0.62 | 1.15 | 1.00 | 157,038 | <0.001 |
| **Week** | 0.03 | 0.01 | 1.03 | 1.01 | 1.04 | 1.00 | 124,639 | <0.001 |
| **Temperature (Z-score)** | -0.11 | 0.04 | 0.90 | 0.84 | 0.96 | 1.00 | 120,296 | <0.001 |

**eTable 2A. Three-way interaction negative binomial regression model (n=1830 games).** The median IRR for the three-way interaction term was 1.0004 (i.e., a trivial effect) and the 50% credible interval crossed 1.0. This supported our *a priori* model design (i.e., it was not necessary to include the three-way interaction in the model). (Note, the IRR for week is reported as 1.00, but is 0.997, so it does cross 1.0).

|  | **Β** | | **Incidence Rate Ratio (IRR)** | **95% Credible Interval for IRR** | | **MCMC Diagnostics** | | |
| --- | --- | --- | --- | --- | --- | --- | --- | --- |
| **Parameter** | **Median** | **SD** | **Median** | **Lower (2.5%)** | **Upper (97.5%)** | **R-hat** | **ESS** | **MCSE** |
| **Grass Surface** | -0.24 | 0.07 | 0.78 | 0.68 | 0.91 | 1.00 | 111,788 | <0.001 |
| **Week** | 0.02 | 0.01 | 1.02 | 1.00 | 1.04 | 1.00 | 81,503 | <0.001 |
| **Temperature (Z-score)** | -0.17 | 0.07 | 0.85 | 0.73 | 0.98 | 1.00 | 59,115 | <0.001 |
| **Grass** × **Week** | 0.02 | 0.02 | 1.02 | 0.99 | 1.05 | 1.00 | 78,537 | <0.001 |
| **Grass** × **Temperature (Z-score)** | 0.10 | 0.09 | 1.11 | 0.93 | 1.32 | 1.00 | 58,215 | <0.001 |
| **Week** × **Temperature (Z-score)** | 0.00 | 0.01 | 1.00 | 0.97 | 1.03 | 1.00 | 62,354 | <0.001 |
| **Grass** × **Week** × **Temperature (Z-score)** | 0.00 | 0.02 | 1.00 | 0.97 | 1.04 | 1.00 | 59,845 | <0.001 |

**eTable 2B. Two-way interaction (all) negative binomial regression model (n=1830 games).** The median IRR for the Week × Temperature interaction term was at 1.0003 (a trivial effect), with a 50% confidence interval which crossed 1.0. This was consistent with our *a priori* model design (i.e., inclusion of this interaction term was not justified in the model). (Note, the IRR for week is reported as 1.00, but is 0.997, so it does cross 1.0).

|  | **β** | | **Incidence Rate Ratio (IRR)** | **95% Credible Interval for IRR** | | **MCMC Diagnostics** | | |
| --- | --- | --- | --- | --- | --- | --- | --- | --- |
| **Parameter** | **Median** | **SD** | **Median** | **Lower (2.5%)** | **Upper (97.5%)** | **R-hat** | **ESS** | **MCSE** |
| **Grass Surface** | -0.25 | 0.07 | 0.78 | 0.68 | 0.89 | 1.00 | 153,603 | <0.001 |
| **Week** | 0.02 | 0.01 | 1.02 | 1.00 | 1.04 | 1.00 | 95,541 | <0.001 |
| **Temperature** | -0.17 | 0.06 | 0.84 | 0.75 | 0.95 | 1.00 | 83,829 | <0.001 |
| **Grass** × **Week** | 0.02 | 0.02 | 1.02 | 0.99 | 1.05 | 1.00 | 96,652 | <0.001 |
| **Grass** × **Temperature (Z-score)** | 0.11 | 0.07 | 1.11 | 0.96 | 1.29 | 1.00 | 88,584 | <0.001 |
| **Week** × **Temperature (Z-score)** | 0.00 | 0.01 | 1.00 | 0.99 | 1.02 | 1.00 | 136,122 | <0.001 |

**eTable 2C. Two-way interaction (all) negative binomial regression model (n=1830 games), using a relaxed prior for the intercept.** The intercept was set to 0 and the scale was set to 0.25. The model was virtually identical to that with the pre-specified priors reported in Table 2 of the main manuscript.

|  | **β** | | **Incidence Rate Ratio (IRR)** | **95% Credible Interval for IRR** | | **MCMC Diagnostics** | | |
| --- | --- | --- | --- | --- | --- | --- | --- | --- |
| **Parameter** | **Median** | **SD** | **Median** | **Lower (2.5%)** | **Upper (97.5%)** | **R-hat** | **ESS** | **MCSE** |
| **Grass Surface** | -0.25 | 0.07 | 0.78 | 0.69 | 0.89 | 1.00 | 148,464 | <0.001 |
| **Week** | 0.02 | 0.01 | 1.02 | 0.997 | 1.04 | 1.00 | 93,743 | <0.001 |
| **Temperature** | 0.09 | 0.06 | 0.85 | 0.76 | 0.95 | 1.00 | 89,884 | <0.001 |
| **Grass** × **Week** | 0.02 | 0.02 | 1.02 | 0.99 | 1.05 | 1.00 | 96,068 | <0.001 |
| **Grass** × **Temperature (Z-score)** | 0.11 | 0.07 | 1.11 | 0.96 | 1.28 | 1.00 | 89,006 | <0.001 |

**eTable 2D. Two-way interaction (all) negative binomial regression model (n=1830 games), using a relaxed prior for the intercept.** The intercept was set to 0 and the scale was set to 1.0. The model was virtually identical to that with the pre-specified priors reported in Table 2 of the main manuscript.

|  | **β** | | **Incidence Rate Ratio (IRR)** | **95% Credible Interval for IRR** | | **MCMC Diagnostics** | | |
| --- | --- | --- | --- | --- | --- | --- | --- | --- |
| **Parameter** | **Median** | **SD** | **Median** | **Lower (2.5%)** | **Upper (97.5%)** | **R-hat** | **ESS** | **MCSE** |
| **Grass Surface** | -0.25 | 0.07 | 0.78 | 0.68 | 0.89 | 1.00 | 147,035 | <0.001 |
| **Week** | 0.02 | 0.01 | 1.02 | 0.997 | 1.04 | 1.00 | 94,780 | <0.001 |
| **Temperature** | -0.17 | 0.06 | 0.85 | 0.76 | 0.95 | 1.00 | 90,157 | <0.001 |
| **Grass** × **Week** | 0.02 | 0.02 | 1.02 | 0.99 | 1.05 | 1.00 | 95,204 | <0.001 |
| **Grass** × **Temperature (Z-score)** | 0.11 | 0.07 | 1.11 | 0.96 | 1.29 | 1.00 | 89,435 | <0.001 |

**eTable 3A. Main effects negative binomial regression model.** If the two-way interaction terms included in the primary model in Table 2 of the manuscript are interpreted as “insignificant” (i.e., probability of effect <90% for each), they may be removed from the model to produce a main effects model. Beyond the 95% credible intervals (reported below), all three main effects have 99% credible intervals which do not cross 1.0, indicating that grass surface, early season games, and warmer temperatures are all associated with a reduced risk of concussion. Visual representations of the marginal effects for this model are presented in eFigures 6A-B and 7A-B.

|  | **β** | | **Incidence Rate Ratio (IRR)** | **95% Credible Interval for IRR** | | **MCMC Diagnostics** | | |
| --- | --- | --- | --- | --- | --- | --- | --- | --- |
| **Parameter** | **Median** | **SD** | **Median** | **Lower (2.5%)** | **Upper (97.5%)** | **R-hat** | **ESS** | **MCSE** |
| **Grass Surface** | -0.26 | 0.06 | 0.77 | 0.68 | 0.88 | 1.00 | 163,734 | <0.001 |
| **Week** | 0.03 | 0.01 | 1.03 | 1.01 | 1.04 | 1.00 | 140,750 | <0.001 |
| **Temperature (Z-score)** | -0.11 | 0.04 | 0.90 | 0.84 | 0.96 | 1.00 | 141,250 | <0.001 |

**eTable 3B. Predicted number of concussions per game for selected game conditions using main effects model.** Results are nearly identical to that using the model with the relevant two-way interactions included.

| **Conditions** | **Grass** | **Artificial Turf** |
| --- | --- | --- |
| **Average week (8-9) with**  **average temperature (~17^o^C)** | 0.52 | 0.67 |
| **Early season (Week 1), with**  **warm weather (25^o^C)** | 0.38 | 0.49 |
| **Late season (Week 16), with**  **cold weather (5^o^C)** | 0.73 | 0.95 |

**eTables 4. Negative binomial regression models using a subset of data at relatively fixed temperature.**

**4A. Only indoor games on artificial turf (n=359**). When only indoor games were examined, there was a 90% probability that week was associated with concussion risk. (Note, the IRR for week is reported as 1.00, but is 0.996, so it does cross 1.0). [Note, there were 63 indoor games played on natural grass, with 25 concussions. These games were excluded from this model to simplify it, and completely remove the effect of surface.]

|  | **Β** | | **Incidence Rate Ratio (IRR)** | **95% Credible Interval for IRR** | | **MCMC Diagnostics** | | |
| --- | --- | --- | --- | --- | --- | --- | --- | --- |
| **Parameter** | **Median** | **SD** | **Median** | **Lower (2.5%)** | **Upper (97.5%)** | **R-hat** | **ESS** | **MCSE** |
| **Week** | 0.03 | 0.02 | 1.03 | 1.00 | 1.06 | 1.000 | 138,034 | <0.001 |

**4B. All games (natural grass + artificial turf) played at a temperature 17.8 to 25.6^o^C (n=827) – interactions included.** When the two way interaction of Surface × Week was included in the model, the probability for an interaction effect was <50%. Thus, it was removed from the model (4C).

|  | **Β** | | **Incidence Rate Ratio (IRR)** | **95% Credible Interval for IRR** | | **MCMC Diagnostics** | | |
| --- | --- | --- | --- | --- | --- | --- | --- | --- |
| **Parameter** | **Median** | **SD** | **Median** | **Lower (2.5%)** | **Upper (97.5%)** | **R-hat** | **ESS** | **MCSE** |
| **Surface** | -0.27 | 0.10 | 0.77 | 0.62 | 0.94 | 1.000 | 159,555 | <0.001 |
| **Week** | 0.02 | 0.01 | 1.02 | 0.99 | 1.05 | 1.000 | 106,755 | <0.001 |
| **Surface X Week** | 0.01 | 0.02 | 1.01 | 0.97 | 1.06 | 1.000 | 106,599 | <0.001 |

**4C. All games (natural grass + artificial turf) played at a temperature 17.8 to 25.6^o^C (n=827) – main effects-only.** In the main effects model, there was a 98% probability that Week was associated with concussion risk, and also a 98% probability that Surface was associated with concussion risk.

|  | **Β** | | **Incidence Rate Ratio (IRR)** | **95% Credible Interval for IRR** | | **MCMC Diagnostics** | | |
| --- | --- | --- | --- | --- | --- | --- | --- | --- |
| **Parameter** | **Median** | **SD** | **Median** | **Lower (2.5%)** | **Upper (97.5%)** | **R-hat** | **ESS** | **MCSE** |
| **Surface** | -0.26 | 0.10 | 0.77 | 0.63 | 0.94 | 1.000 | 172,915 | <0.001 |
| **Week** | 0.03 | 0.01 | 1.03 | 1.01 | 1.05 | 1.000 | 176,644 | <0.001 |

**eTables 5. Negative binomial regression models using a subset of late season games (weeks 13-16).** The wide credible intervals for the two-way interactions suggested a weak effect (5A), thus they were removed from the model (5B).

**5A. Model with two-way interactions.** When the two-way interactions were included (without week included in the model), there was a 95% probability that temperature was associated with concussion risk. Note, the 97.5% credible interval is 0.997, and thus does not cross 1.00.

|  | **β** | | **Incident Rate Ratio (IRR)** | **95% Credible Interval for IRR** | | **MCMC Diagnostics** | | |
| --- | --- | --- | --- | --- | --- | --- | --- | --- |
| **Parameter** | **Median** | **SD** | **Median** | **Lower (2.5%)** | **Upper (97.5%)** | **R-hat** | **ESS** | **MCSE** |
| **Grass Surface** | -0.28 | 0.12 | 0.76 | 0.60 | 0.96 | 1.000 | 120,866 | <0.001 |
| **Temperature (Z-score)** | -0.17 | 0.09 | 0.84 | 0.71 | 1.00 | 1.000 | 83,949 | <0.001 |
| **Grass** × **Temperature (Z-score)** | 0.13 | 0.12 | 1.14 | 0.91 | 1.43 | 1.000 | 85,260 | <0.001 |

**5B. Main effects-only model without week.** In this model, there was a 91% probability that temperature influenced concussion risk, a 98% probability grass surface influenced concussion risk.

|  | **Β** | | **Incidence Rate Ratio (IRR)** | **95% Credible Interval for IRR** | | **MCMC Diagnostics** | | |
| --- | --- | --- | --- | --- | --- | --- | --- | --- |
| **Parameter** | **Median** | **SD** | **Median** | **Lower (2.5%)** | **Upper (97.5%)** | **R-hat** | **ESS** | **MCSE** |
| **Grass Surface** | -0.29 | 0.12 | 0.75 | 0.60 | 0.94 | 1.000 | 175,011 | <0.001 |
| **Temperature (Z-score)** | -0.10 | 0.06 | 0.90 | 0.81 | 1.01 | 1.000 | 177,514 | <0.001 |

**eTable 6. Summary of regression parameters when natural grass was compared to Field Turf brand artificial surface.** Results are similar to the primary model, which included all brands of artificial turf. The two-way interaction terms demonstrated a very weak effect (6A) and thus were removed from the model (6B).

**6A. Relevant two-way interaction terms included.**

|  | **Β** | | **Incidence Rate Ratio (IRR)** | **95% Credible Interval for IRR** | | **MCMC Diagnostics** | | |
| --- | --- | --- | --- | --- | --- | --- | --- | --- |
| **Parameter** | **Median** | **SD** | **Median** | **Lower (2.5%)** | **Upper (97.5%)** | **R-hat** | **ESS** | **MCSE** |
| **Grass Surface** | -0.23 | 0.09 | 0.79 | 0.66 | 0.96 | 1.000 | 112,786 | <0.001 |
| **Week** | 0.03 | 0.02 | 1.03 | 0.99 | 1.07 | 1.000 | 67,040 | <0.001 |
| **Temperature (Z-score)** | -0.23 | 0.09 | 0.80 | 0.66 | 0.96 | 1.000 | 65,440 | <0.001 |
| **Grass** × **Week** | 0.00 | 0.02 | 1.01 | 0.97 | 1.05 | 1.000 | 66,742 | <0.001 |
| **Grass** × **Temperature (Z-score)** | 0.16 | 0.11 | 1.17 | 0.95 | 1.45 | 1.000 | 65,437 | <0.001 |

**eTable 6B. Main effects-only model.** When the two-way interaction terms from 6A are removed, all main effects parameters have 95% credible intervals which do not cross 1.0.

|  | **β** | | **Incidence Rate Ratio (IRR)** | **95% Credible Interval for IRR** | | **MCMC Diagnostics** | | |
| --- | --- | --- | --- | --- | --- | --- | --- | --- |
| **Parameter** | **Median** | **SD** | **Median** | **Lower (2.5%)** | **Upper (97.5%)** | **R-hat** | **ESS** | **MCSE** |
| **Grass Surface** | -0.27 | 0.09 | 0.76 | 0.64 | 0.91 | 1.000 | 164,115 | <0.001 |
| **Week** | 0.03 | 0.01 | 1.04 | 1.02 | 1.06 | 1.000 | 123,253 | <0.001 |
| **Temperature (Z-score)** | -0.10 | 0.04 | 0.91 | 0.83 | 0.99 | 1.000 | 123,262 | <0.001 |

**eFigure 1. Posterior distribution of surface parameter estimates in the preliminary main effects model, including games played on hybrid surfaces.** Shaded areas represent the Bayesian 89% credible interval and tails represent that Bayesian 97% credible interval. The dark grey vertical bar represents the median Incidence rate ratio (IRR). IRR <1.0 indicate reduced risk of concussion on natural grass compared to artificial turf (reference category). Hybrid surface may also be associated with a reduced risk of concussions compared to artificial turf, but the probability was only 69% (as evidenced by the wide 89% credible interval). Conversely, the probability for natural grass to be associated with a reduced risk of concussion compared to artificial turf was >99%. Given the clear effect of natural grass, and the possible, but less certain effect of hybrid surface on concussion risk, combined with the small sample size, games played on hybrid surface was excluded from all other analyses.

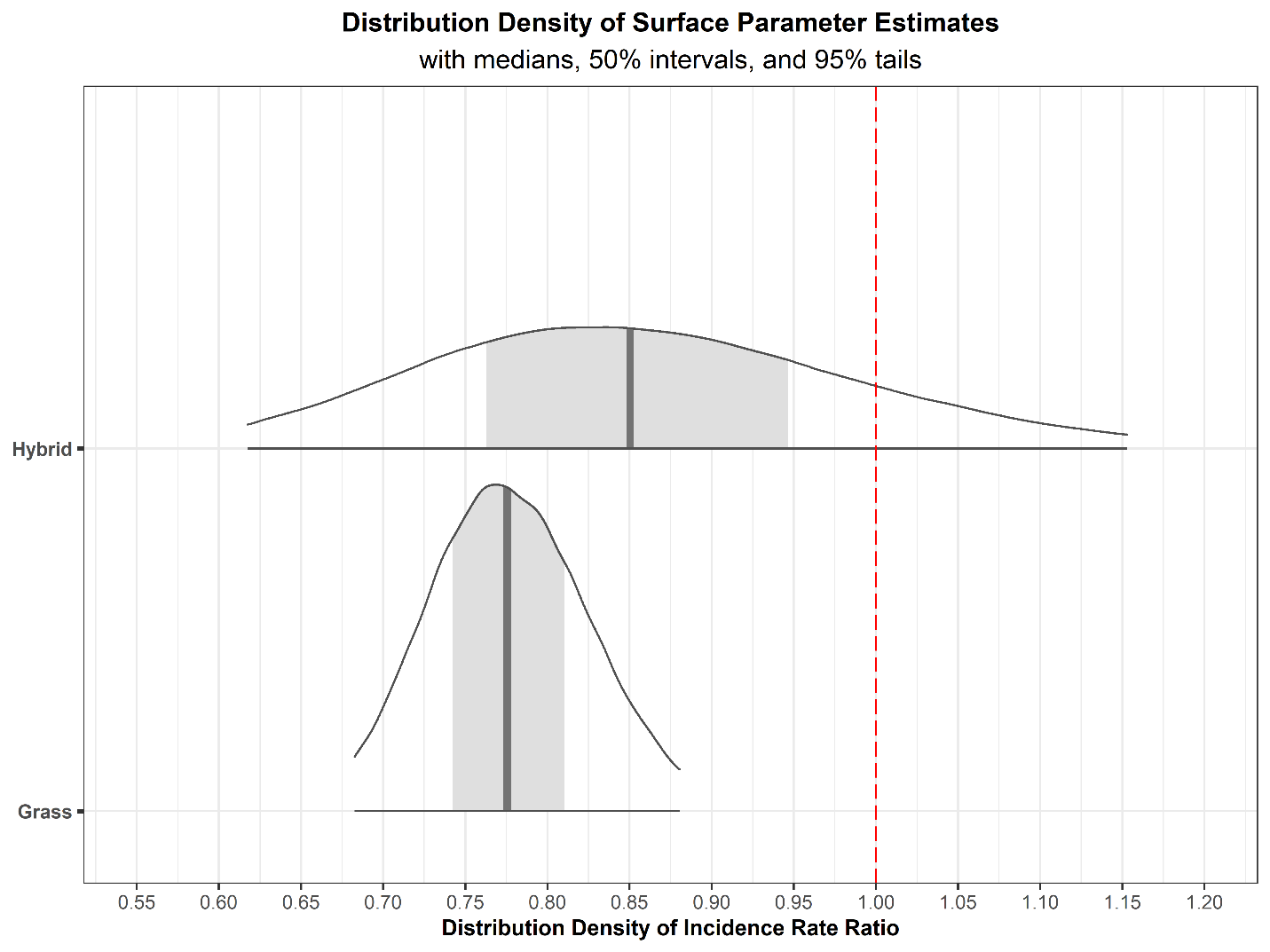

**eFigure 2. Histogram of the number of concussions per game across 8 NFL seasons.**

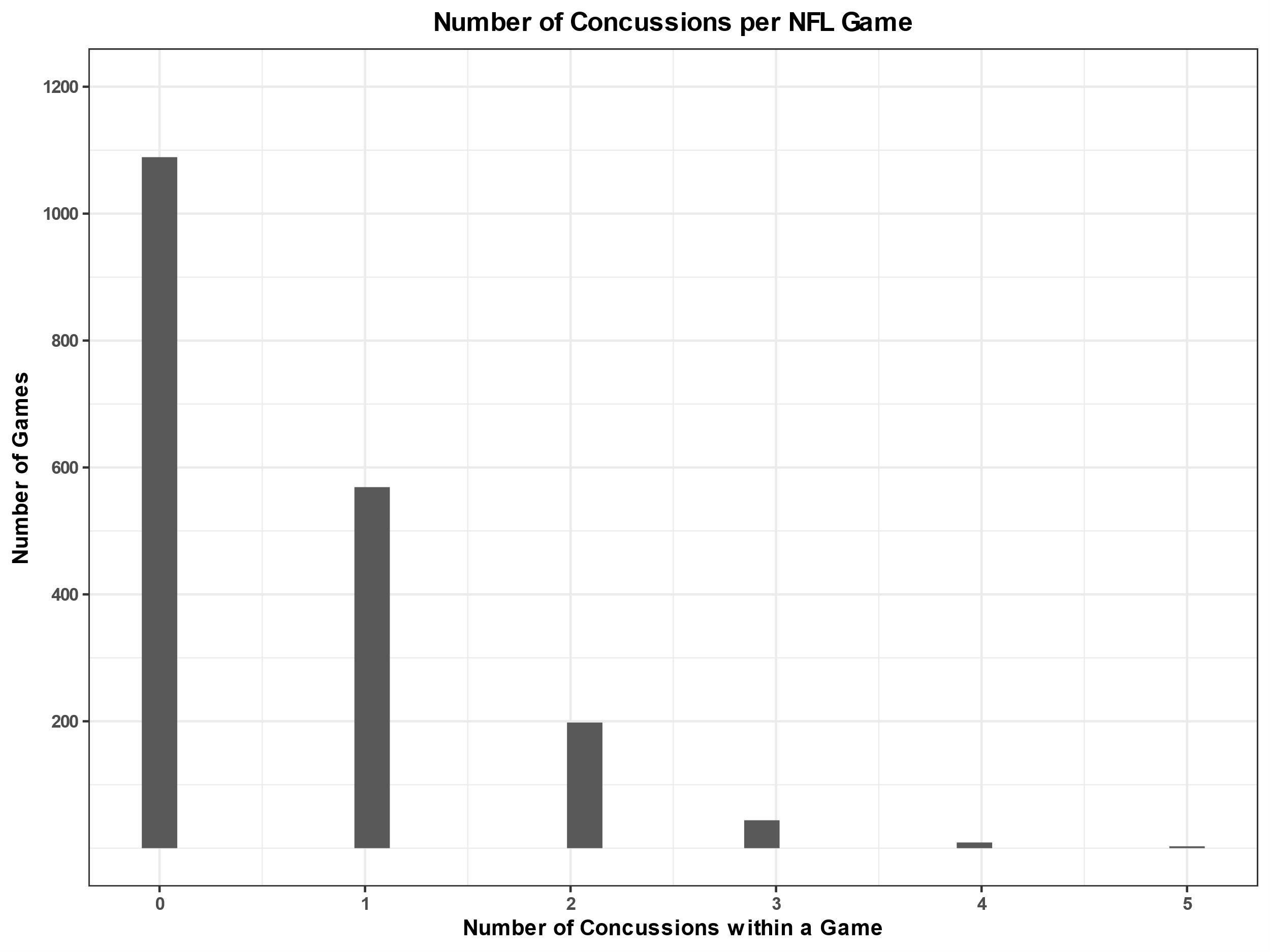

**eFigure 3. Distribution of temperature by surface and by week.** The black line on 3A and 3B represents median temperature. Note, the spike at 20^o^C is due to numerous games on artificial turf played indoors. For outdoor games, the lower and upper quartile temperatures were 7.8 and 18.9^o^C for artificial turf and 12.2 and 24.4^o^C for natural grass, respectively.

**3A. All games (indoor + outdoor) on natural grass.**

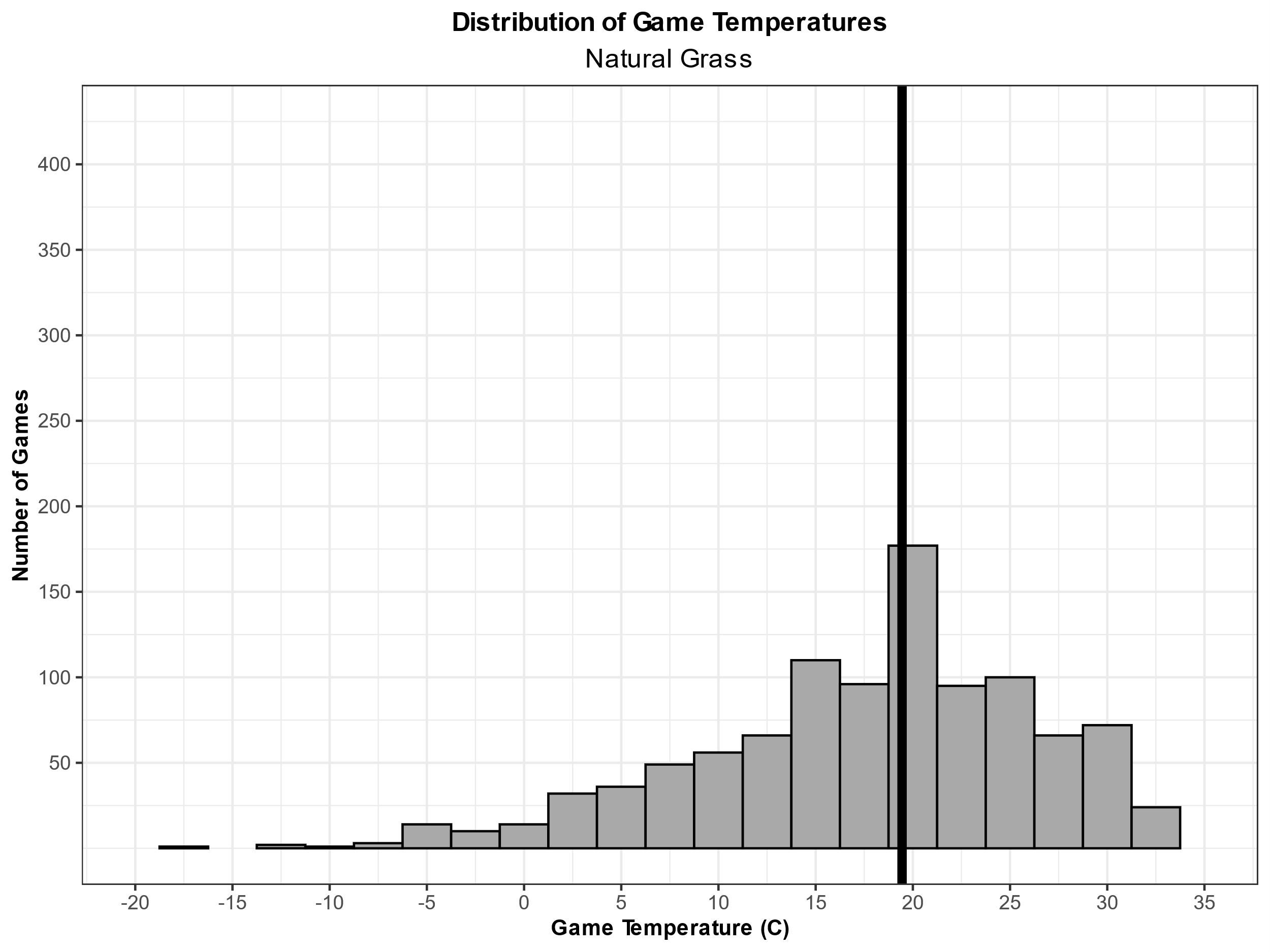

**3B. All games (indoor + outdoor) on artificial turf.**

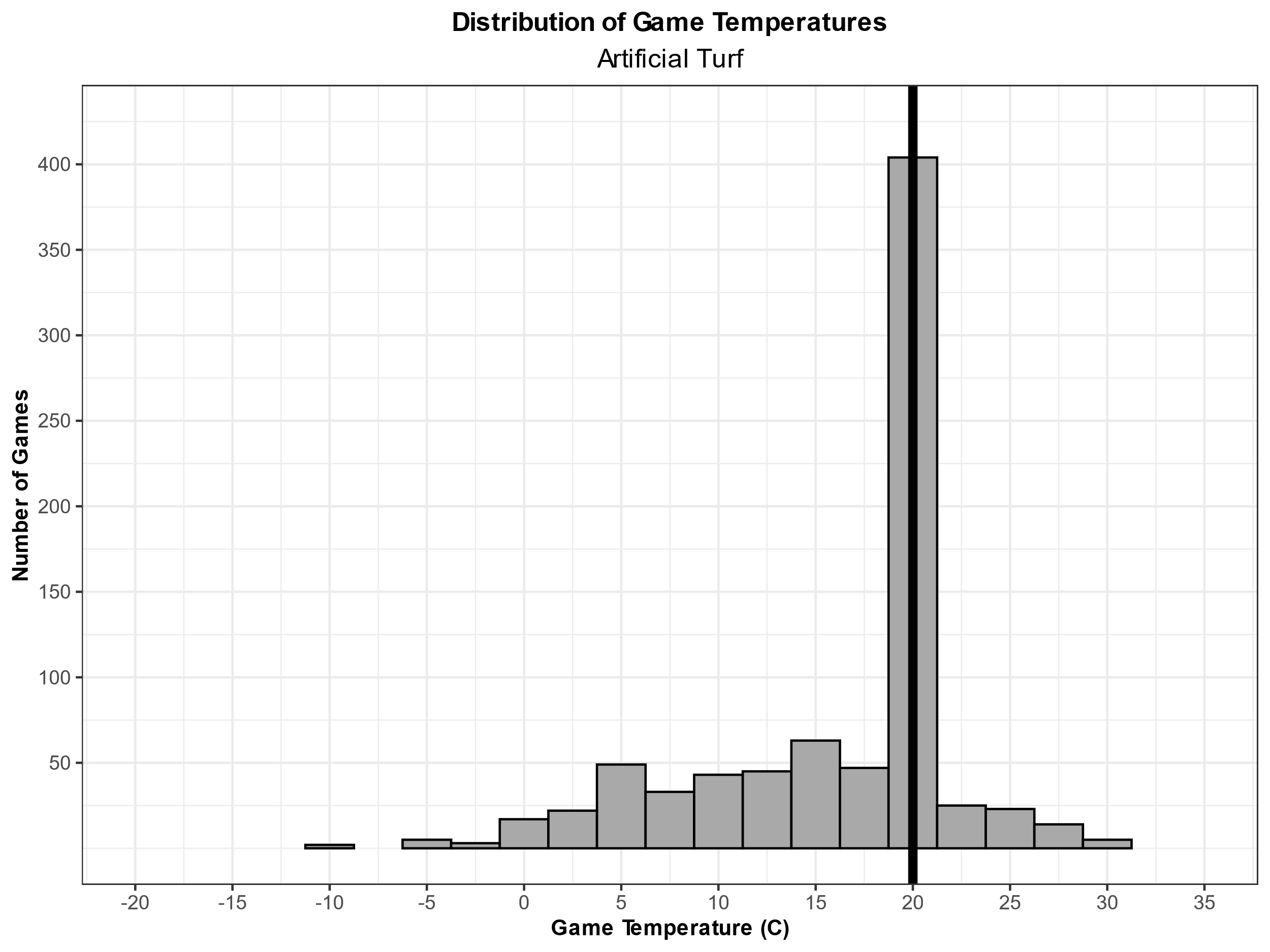

**3C. Comparison of Temperature by Week of NFL Season.**

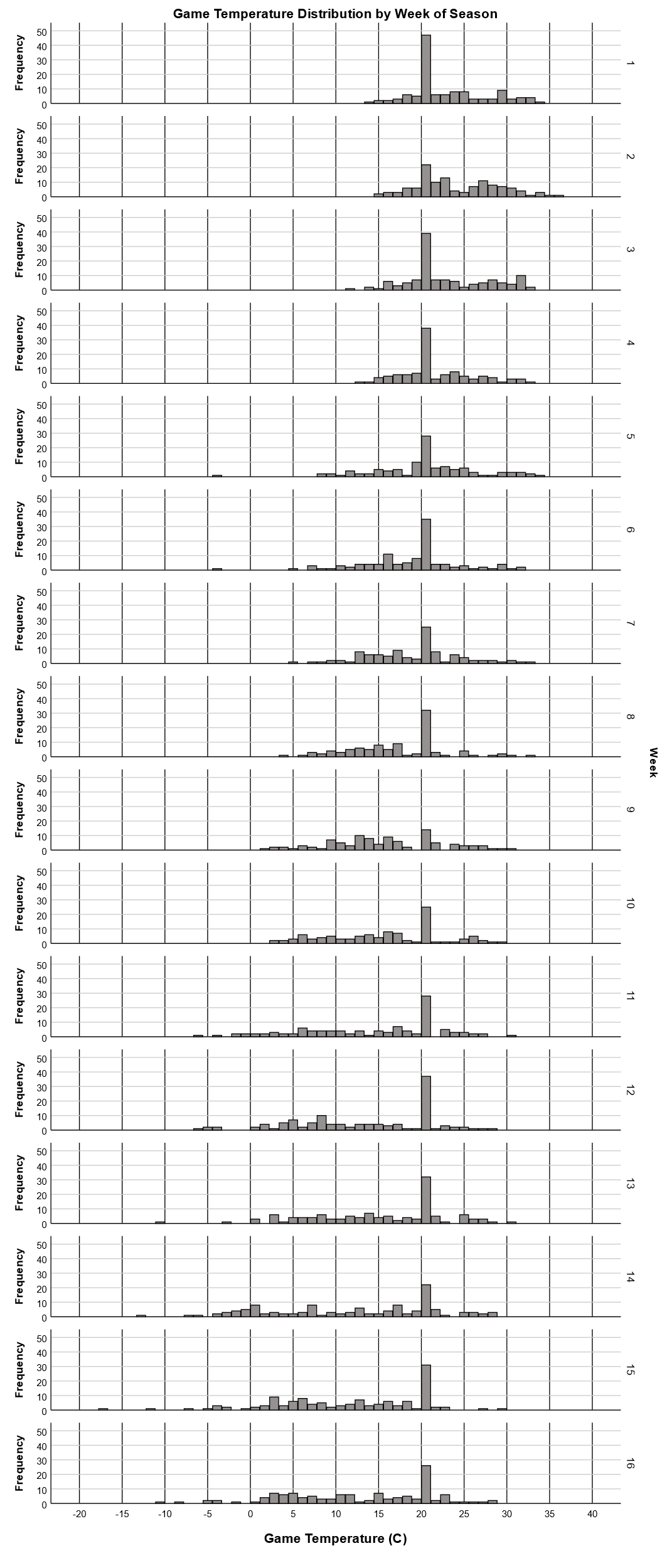

**eFigure 4. Monte Carlo Markov Chain (MCMC) trace plots for each parameter in the model.** All parameters appeared to reach convergence for each chain.

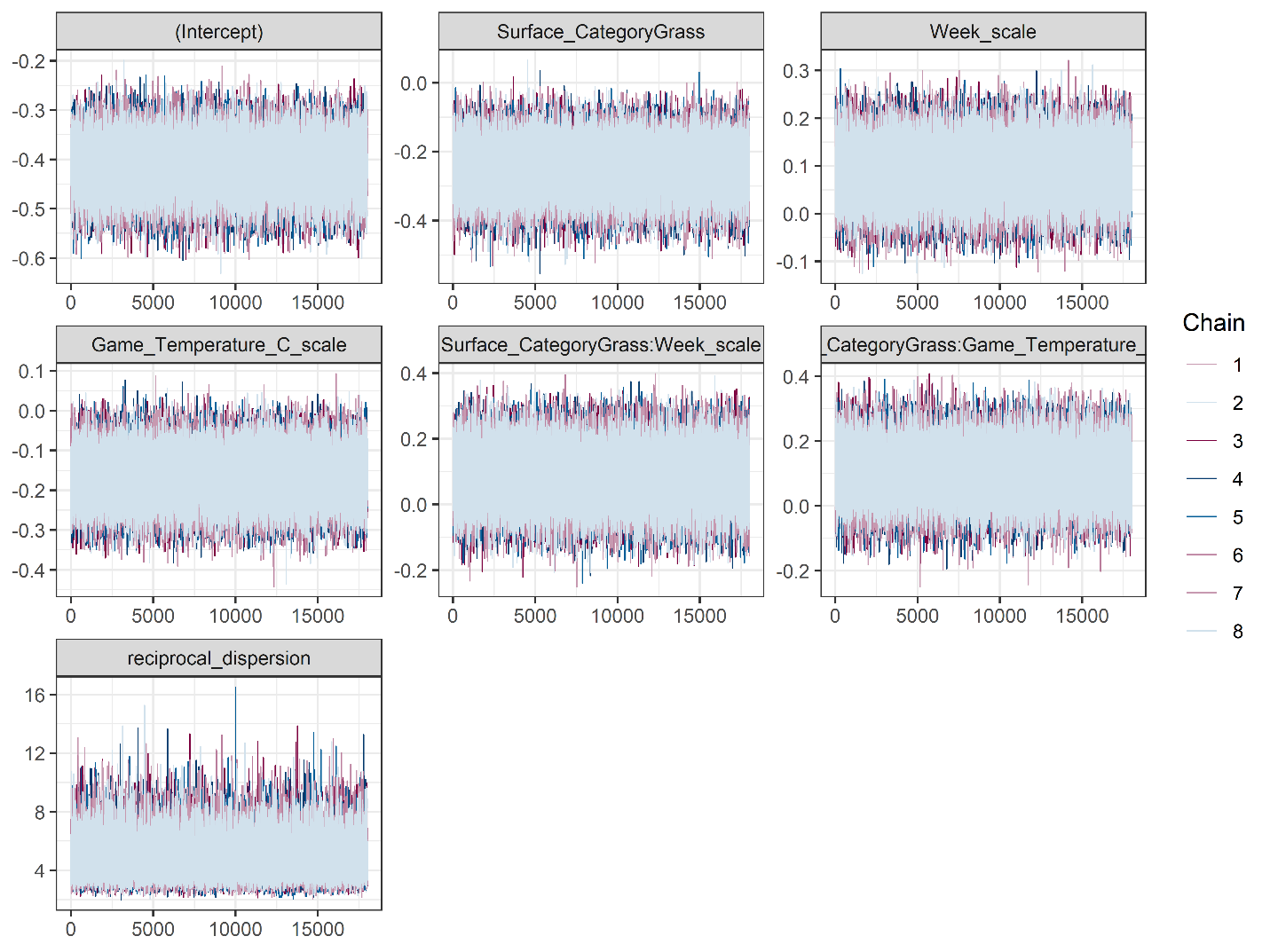

**eFigure 5.** **Observed vs. model-predicted counts of concussion across NFL games.** The black lines represent observed data, and the grey lines (which coalesce to form shading) represent 1000 simulations based on the model’s posterior predictive distributions. The close relationship between observed and predicted data indicate an excellent model fit.

**5A. Natural grass and artificial turf combined.**

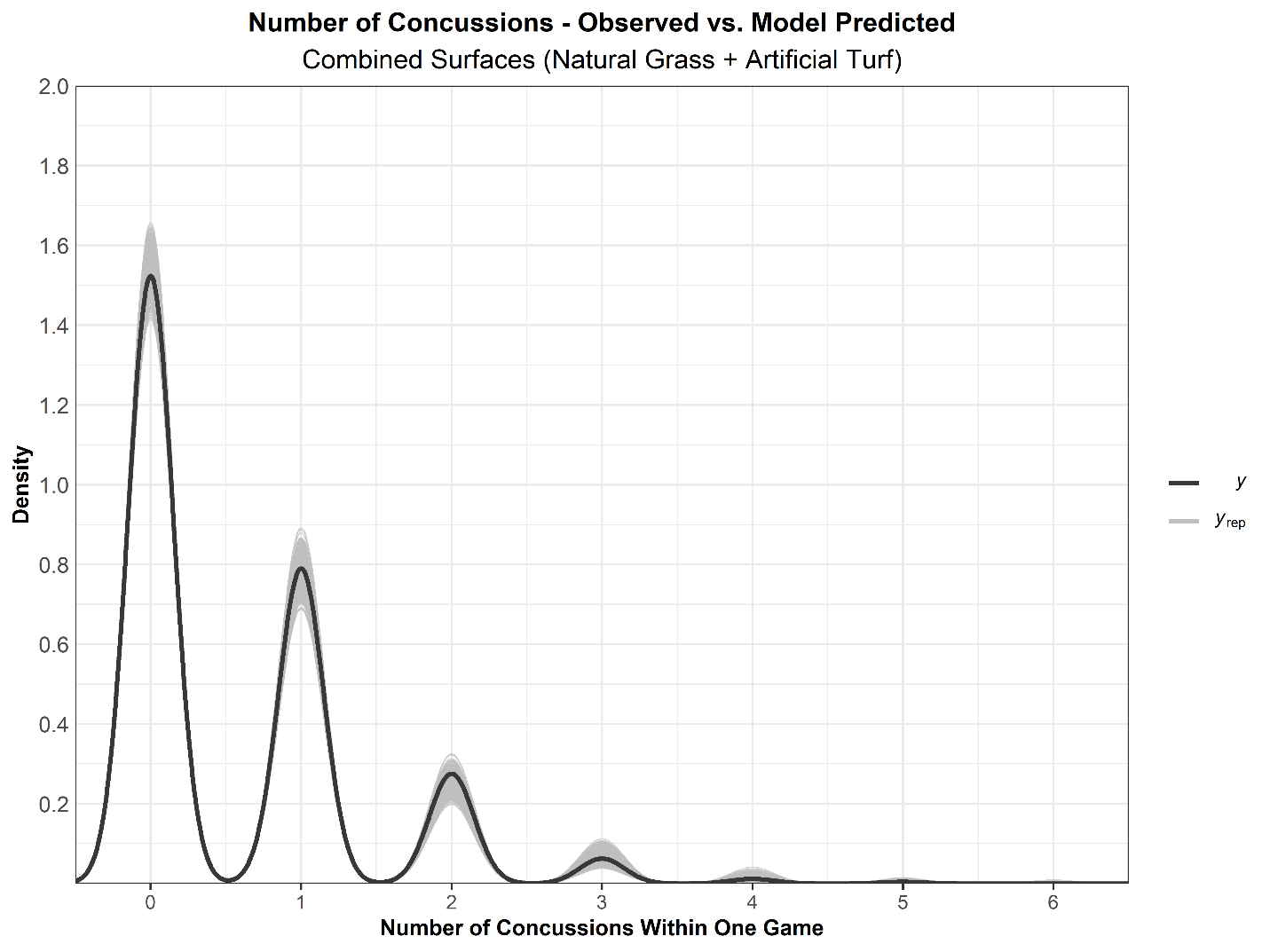

**5B. Natural grass and artificial turf separated.**

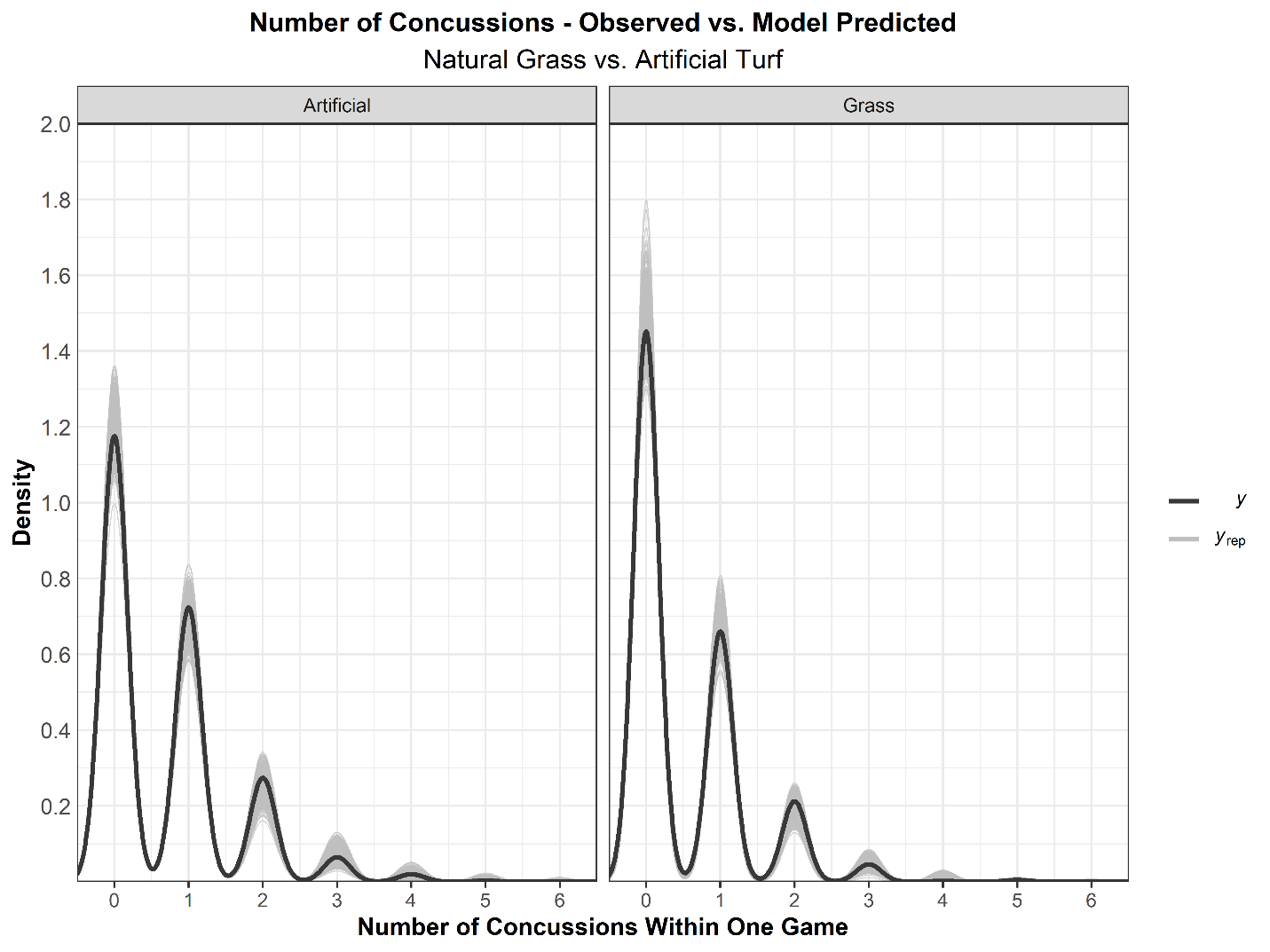

**eFigure 6. Marginal effects of concussion rate by week of season at fixed temperatures, using main effects only model.** If the two-way interaction terms are removed from the model, there remain clear effects for surface, week, and temperature (eTable 3). However, the reduced risk of concussion on natural grass compared to artificial turf is magnified in the main effects-only model at 20C and 25C and slightly attenuated at 15C (see Figure 2 of main manuscript for comparison to model with relevant interaction terms).

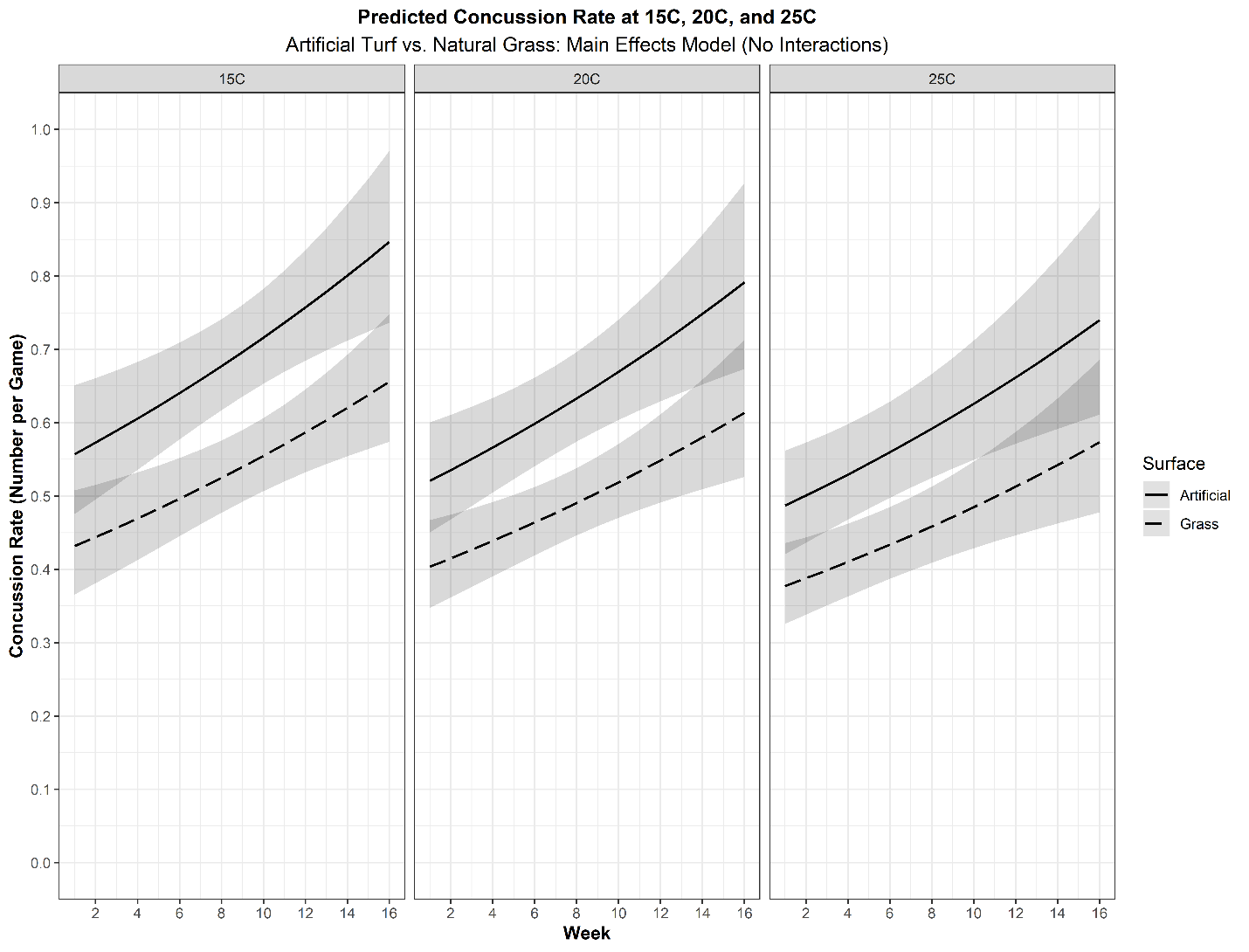

**eFigure 7. Marginal effects of concussion rate by week of season at fixed week of the season, using main effects only model.** If the two-way interaction terms are removed from the model (eTable 3), the results are similar to that with the interaction terms included (Table 2, main manuscript). At low temperature, the reduced risk of concussion on natural grass compared to artificial turf is attenuated (yet still present) in the main effects-only model. However, the main effects model also exacerbates the difference in risk between surfaces at higher temperatures. As described in Figure 3 of the main manuscript, the mean temperature for outdoor games at Week 9 is 15^o^C. The red and blue vertical lines represent the observed game temperature extremes at Week 9 for artificial turf and natural grass, respectively.

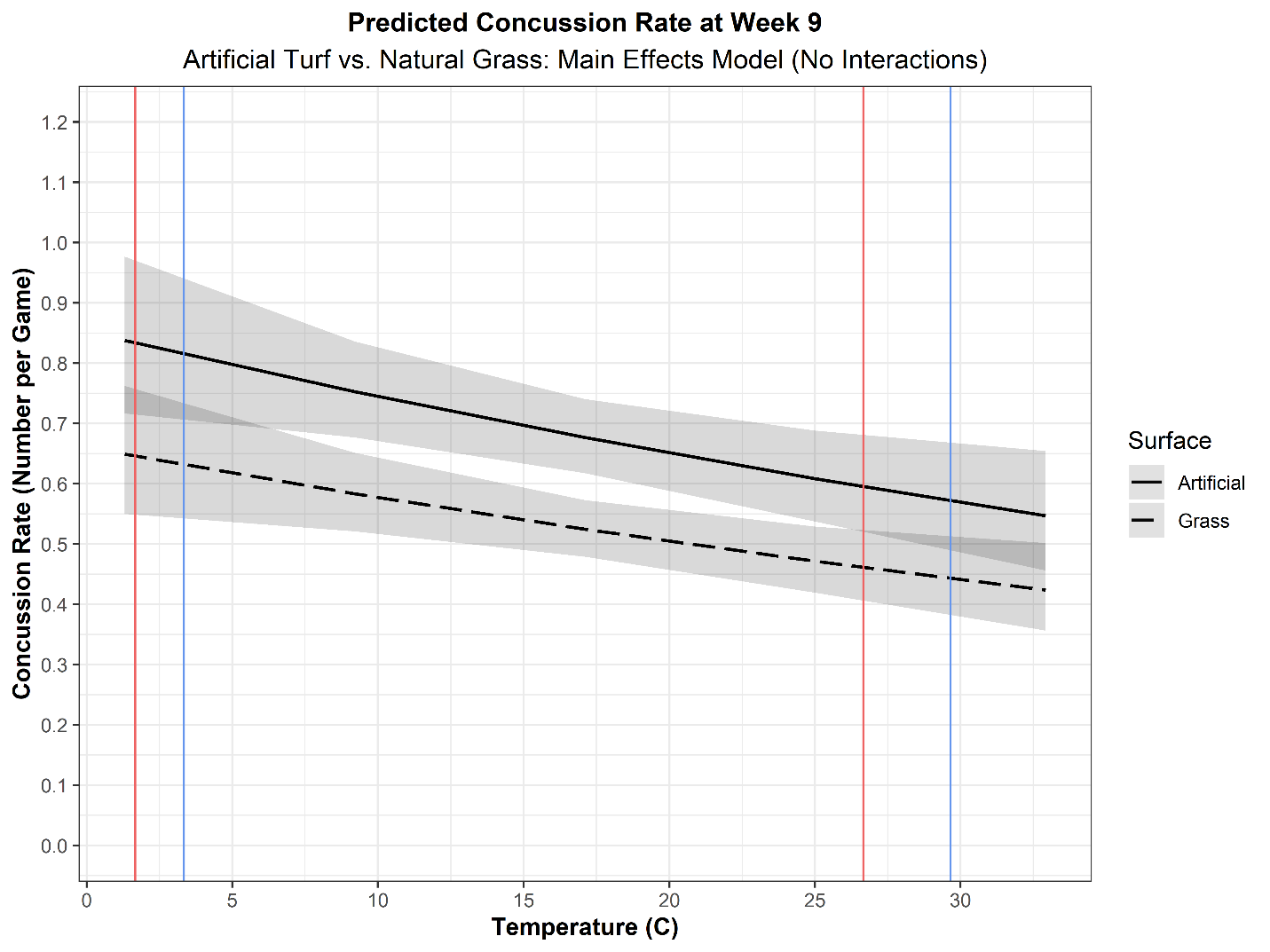

**eFigure8.** **Marginal effects of concussion rate comparing natural grass to FieldTurf artificial surfaces using main effects-only model (see Table 6B).** We compared natural grass to this one specific brand of artificial turf, since previous industry-funded research reported that risk of concussion was reduced on FieldTurf compared to natural grass in college football ^2^. Of all games played on artificial turf (n=800), 37.9% (n=303) were played on FieldTurf. smaller sample size is likely responsible for the relatively greater overlap of the credible intervals, compared to those in Figure 2 and Figure 3 of the main text. The mean temperatures of games played on FieldTurf and natural grass at were 15.7 ^o^C and 17.7^o^C, respectively.

**eFigure 8A. Marginal effects of concussion rate by week of season at fixed temperatures, using main effects only model, comparing natural grass to FieldTurf.** If the two-way interaction terms are removed from the model, there remain clear effects for surface, week, and temperature (eTable 6B). There is clear separation (i.e. no overlap in credible interval bands) at cooler temperatures during the middle of the season.

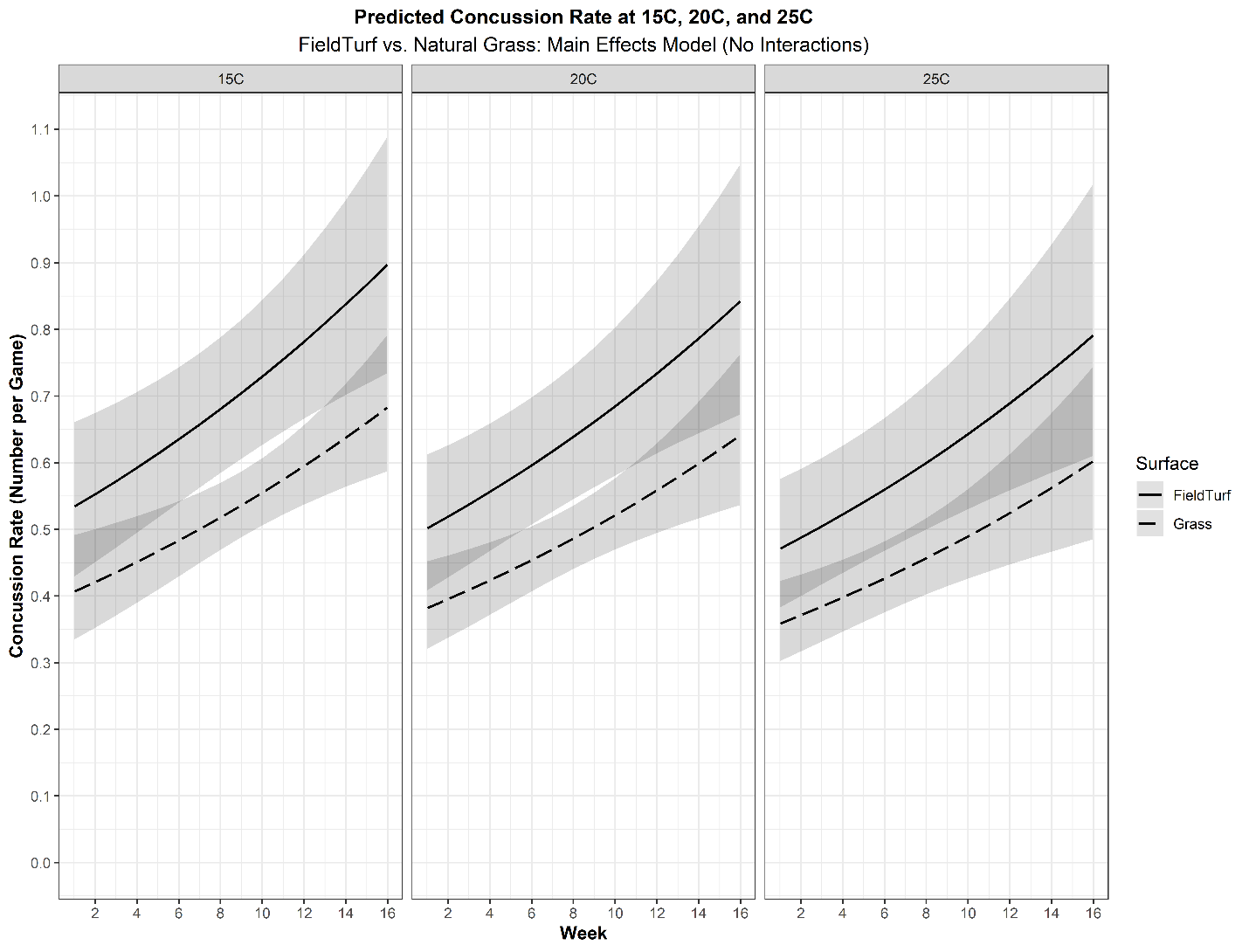

**eFigure 8B. Marginal effects of concussion rate by temperature, fixed to Week 9 of the NFL season, comparing natural grass to FieldTurf.** There is substantial overlap in the 95% credible intervals at extreme temperatures, but this is likely attributable to lack of data at these extremes. Red lines represent the maximum and minimum temperature observed at Week 9 for FieldTurf. Blue lines represent the maximum and minimum temperatures observed at Week 9 for Natural Grass. At Week 9, the mean temperature on FieldTurf was 10.8^o^C, and the mean temperature on natural grass was 17.4^o^C.

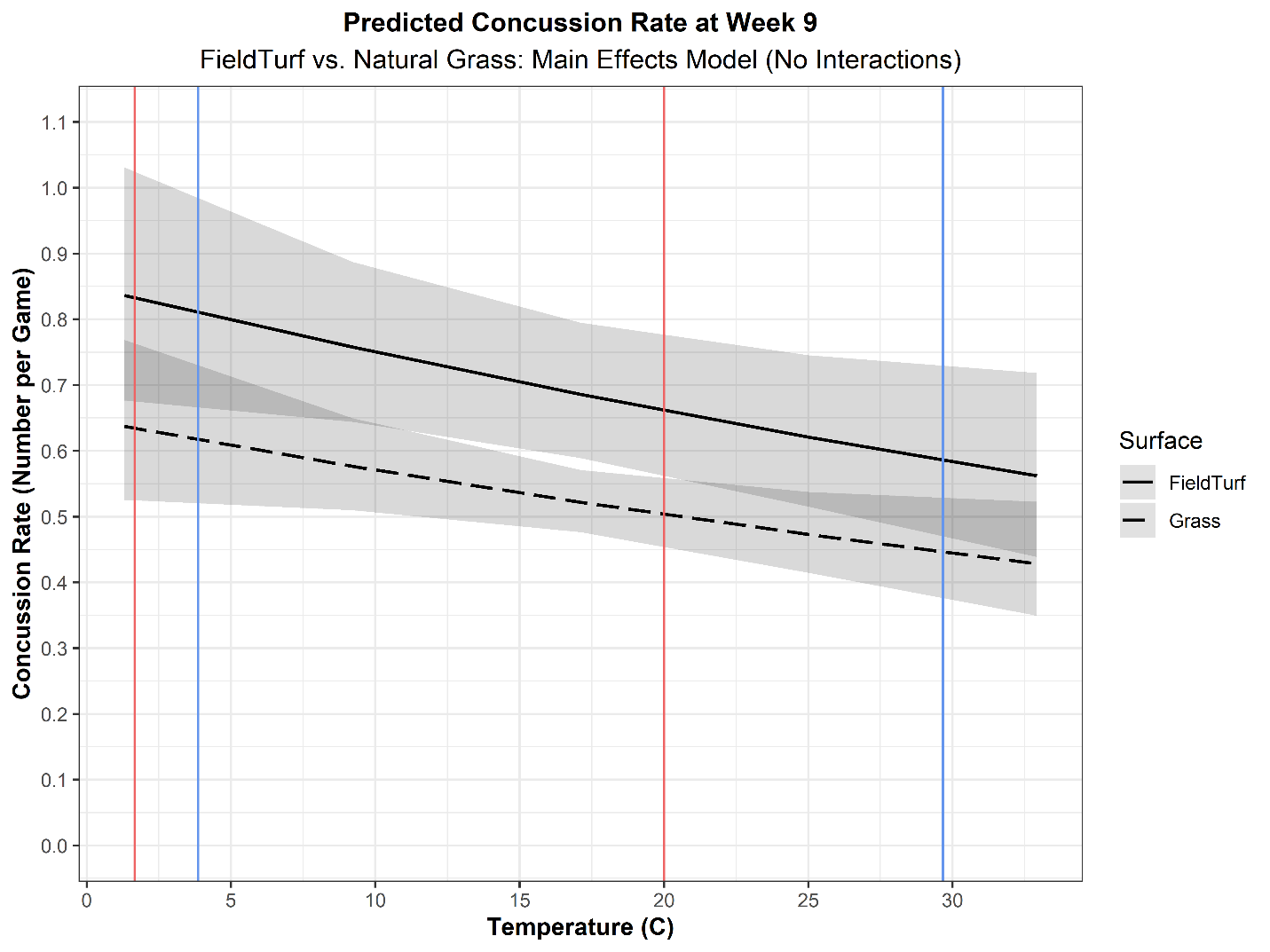

1.	*loo: Efficient leave-one-out cross-validation and WAIC for Bayesian models.* [computer program]. R package version 2.4.1; 2020.

2.	Meyers MC. Incidence, mechanisms, and severity of game-related college football injuries on FieldTurf versus natural grass: a 3-year prospective study. *Am J Sports Med.* 2010;38(4):687-697.
